# Neurovascular instability, impaired cortical recruitment, and network dysconnectivity across the transdiagnostic anxiety spectrum: a functional multi-channel near-infrared spectroscopy study

**DOI:** 10.64898/2026.06.11.26355427

**Authors:** Yiping Luo, Hongzhou Wu, Xia Deng, Luyao Wang, Andre F Carvalho, Yingqian Zhang, Xu Zhang, Michael Maes

## Abstract

**Background:** Anxiety-spectrum disorders (ANSD) are highly prevalent, yet the underlying neurovascular mechanisms remain unclear. Functional near-infrared spectroscopy (fNIRS) comprises a non-invasive method to assess cortical hemodynamics, neurovascular coupling, and network organization during cognitive processing.

**Methods:** We investigated healthy controls (HC), generalized anxiety disorder (GAD), anxious depression (AD), and anxiety–depression comorbidity (CO) using multichannel fNIRS during a verbal fluency task. Multiple hemodynamic features were extracted, including peak response, temporal hemodynamic variability, β₁ activation, and HbO, HbR, and HbT signals. Functional connectivity, graph-theoretical network measures, machine-learning classification, and associations with depressive, anxiety and psychosomatic scores were examined.

**Results:** Compared to controls, ANSD patients showed reduced task-evoked HbO and HbT responses, preserved HbR levels, increased temporal hemodynamic variability, and reduced β₁ activation. Activation deficits were most prominent in bilateral frontopolar and medial prefrontal cortices and followed a gradient, with the CO group exhibiting highest abnormalities. Functional connectivity was increased, whereas clustering coefficient, nodal local efficiency, and nodal efficiency were reduced, indicating maladaptive hyperconnectivity accompanied by inefficient network organization. The AD and CO groups showed the greatest network disintegration. Temporal hemodynamic variability emerged as the strongest predictor of anxiety, depressive, and physiosomatic symptom severity. Reduced prefrontal activation was significantly associated with higher symptom domain scores. Machine-learning analyses demonstrated adequate discrimination between HC and ANSD.

**Conclusions:** ANSD are characterized by impaired neurovascular recruitment, increased hemodynamic instability, maladaptive hyperconnectivity, and disrupted cortical network topology. These abnormalities appear to represent transdiagnostic neurovascular processes underlying anxiety, depressive, and physiosomatic symptoms across the anxiety spectrum.

**Highlights:**

- A 53-channel fNIRS verbal-fluency paradigm revealed widespread neurovascular and cortical network abnormalities across the anxiety spectrum.
- ANSD were characterized by increased hemodynamic variability and reduced peak neurovascular recruitment, indicating unstable yet hyporesponsive cortical dynamics.
- Difference metrics within the right frontal eye field, right Broca’s area, and right frontopolar cortex emerged as robust discriminative biomarkers across machine learning models.
- ANSD exhibited reduced prefrontal activation, impaired network efficiency, and maladaptive hyperconnectivity despite preserved HbR responses.

## Introduction

Anxiety-spectrum disorders (ANSD) comprise a group of common psychiatric conditions characterized by excessive, persistent, and difficult-to-treat anxiety, fear, worry, and avoidance-related behaviors^[1]^. Clinically, this spectrum can range from generalized anxiety disorder (GAD) presentations to comorbid anxiety-depression (CO) and anxious depression (AD)^[2,3]^. Epidemiological evidence suggests that approximately 4.0% of the global population is affected by anxiety disorders, and these conditions impose a substantial public-health burden through somatic symptoms, depressive symptoms, impaired social functioning, health-care use, disability, and increased suicide risk^[4–6]^.Current psychiatric diagnosis and treatment systems still rely heavily on DSM-5- or ICD-11-based structured interviews and subjective symptom-rating scales^[1,7]^.Although these approaches remain indispensable for clinical diagnosis, they are influenced by clinician experience, patient cognitive bias, and expressive ability, and they provide limited direct access to central nervous system pathology^[8]^. Objective biomarkers are therefore urgently needed to support precision psychiatry and individualized treatments^[9,10]^. Neuroimaging studies using fMRI and PET have identified abnormal activation and network alterations in regions such as the frontopolar area (FPA), dorsolateral prefrontal cortex (DLPFC), and Broca’s area in anxiety-spectrum populations^[11–13]^. Increasing physiological evidence further suggests that these macroscopic network abnormalities may reflect local cortical microcirculatory imbalance and impaired neurovascular coupling (NVC)^[14,15]^.

Functional near-infrared spectroscopy (fNIRS) is a non-invasive optical neuroimaging technique based on light penetration through brain tissue and wavelength-specific absorption. It can dynamically monitor cortical hemodynamic changes and estimate relative concentration changes in oxygenated hemoglobin (HbO), deoxygenated hemoglobin (HbR), and total hemoglobin (HbT) across brain regions^[16,17]^. These multidimensional biochemical indicators provide a direct window into NVC and microcirculatory metabolic function^[18]^. Recent neurophysiological evidence suggests that different hemodynamic response patterns may carry distinct pathological implications. For example, reduced HbO accompanied by increased HbT may indicate inefficient NVC, whereby compensatory increases in cerebral blood volume (CBV) fail to adequately support cortical oxygenation demands, reflecting dysregulated microvascular recruitment and impaired metabolic–vascular coupling^[15,17]^. In contrast, reduced HbO with relatively stable HbR may reflect insufficient perfusion or impaired vasomotor responsiveness, indicating blunted neurovascular recruitment^[19,20]^.

Recent studies suggest that fNIRS has potential utility for neurofunctional assessment in anxiety-related disorders. In particular, task-evoked prefrontal hemodynamic abnormalities may help distinguish anxiety disorders, depressive disorders, their comorbidity, and healthy controls^[21–24]^. Given the high comorbidity between anxiety and depression and their overlap in prefrontal regulation, NVC, and microcirculatory dysfunction, it is important to examine GAD, the comorbidity, and anxious depression within a continuous anxiety-spectrum framework^[24,25]^. However, most fNIRS studies remain constrained by traditional diagnostic categories and typically investigate GAD, major depressive disorder (MDD), or their comorbidities as separate entities^[22–24]^. Few studies have integrated clinical phenotyping, multidimensional fNIRS features, and machine-learning approaches to investigate shared abnormalities across the anxiety spectrum^[26,27]^. Consequently, it remains unclear to what extent these fNIRS features map the complex clinical phenotypic variance and disease severity across the anxiety spectrum^[28,29]^.

Furthermore, most previous fNIRS studies focused primarily on a single HbO indicator and rarely integrated HbO, HbR, and HbT to characterize the full profile of cortical microcirculatory hemodynamics^[30,31]^. In feature extraction, conventional analyses typically calculate only the mean signal signs during the task period as an index of functional activation, thereby overlooking the rich temporal, dynamic, and amplitude-related information embedded within hemodynamic signals^[32]^. Therefore, to quantify more comprehensively the dynamic properties of NVC and cortical microcirculatory metabolism, it is more appropriate to systematically extract amplitude-based measures (mean, peak, or integral metrics) and temporal variations (signal fluctuations), and the temporal rate of hemodynamic change (slope metric)^[21,32]^. These dynamic features may improve the accuracy of machine-learning-based disease classification^[33]^ while providing a more better characterization of task-related dysfunction within key cortical regions^[34]^.

Hence, the present study examined high-channel fNIRS data across the anxiety spectrum. First, we compared ANSD with controls using fNIRS-derived features under cognitive load. Second, we examined associations between these multidimensional hemodynamic features and ratings of anxiety, depression, physiosomatic symptoms and overall symptom severity. Third, machine learning models were applied to evaluate the discriminative value of fNIRS assessments among GAD, anxious depression, and comorbid anxiety-MDD. Finally, across the anxiety spectrum, we characterized alterations in activation, functional connectivity, and network topology.

## Materials and Methods

### Participants and study design

A case-control cross-sectional design was adopted. Participants were recruited from the Psychosomatic Medicine Center of Sichuan Provincial People’s Hospital in Chengdu, China. Initially, 268 right-handed participants were enrolled, including 67 HC and 201 inpatients within the transdiagnostic anxiety spectrum, with 67 participants each in the GAD, CO, and AD groups. All patients in the disease groups were diagnosed by a senior psychiatrist through structured clinical interviews according to the Diagnostic and Statistical Manual of Mental Disorders, Fifth Edition (DSM-5). The patients fulfilled DSM-5 diagnostic criteria for the clinical condition assigned through structured clinical interviews and being operationally classified as GAD or MDD, their comorbidity (GAD+MDD, labeled CO) or high anxiety symptoms not fulfilling GAD criteria + MDD (in DSM-5 terms: MDD with anxious distress, here labeled as anxious depression or AD). Other inclusion criteria for patients were as follows: (a) aged between 18 and 65 years; (b) confirmed right-handedness assessed via the Edinburgh Handedness Inventory; (c) having completed at least primary education (≥ 6 years of formal education) to ensure sufficient comprehension for cognitive assessments; and (d) exhibiting no clinically significant structural abnormalities on cranial MRI scans. The healthy control (HC) group consisted of community volunteers and hospital staff, group-matched to participants with ANSD regarding core demographic baselines, including age, gender distribution, years of education, and body mass index (BMI). HCs were required to have no lifetime history of any DSM-5 psychiatric disorders and no family history of MDD, bipolar disorder, substance use disorders, or suicide.

We excluded participants with the following conditions: (a) diagnosis of other major psychiatric or neurodevelopmental disorders (e.g., schizophrenia, bipolar disorder, organic mental disorders, schizoaffective disorder, eating disorders, or autism spectrum disorder); (b) neurological disorders or history of brain injury (e.g., epilepsy, stroke, multiple sclerosis, Alzheimer’s disease, Parkinson’s disease, brain tumors, or traumatic brain injury); (c) severe underlying cardiovascular, hepatic, renal, or endocrine diseases; (d) severe systemic, immunological, or inflammatory diseases (e.g., autoimmune diseases, cancer, or recent severe infections); (e) pregnant or lactating women; (f) history of non-medical drug, alcohol, or substance dependence/abuse (excluding nicotine dependence); and (g) severe cognitive impairment or contraindications for functional near-infrared spectroscopy (fNIRS) testing that precluded task compliance.

All participants or their legal representatives provided written informed consent after a thorough explanation of the study’s objectives, procedures, and potential risks. The study protocol was formally approved by the Ethics Committee of Sichuan Provincial People’s Hospital [Approval No. 331 (2022)].

Using G*Power 3.1.9.4 for a linear multiple regression fixed-model R^2^ deviation from zero test, with f2 = 0.176 (15.0% explained variance), a two-sided alpha level of 0.05, statistical power of 0.80, and up to seven covariates, the recalculated minimum required sample size was 89 participants. The achieved sample size exceeded this requirement, to increase accuracy of the parameter estimates and provide adequate power for multivariate estimation and exploratory train/test validation.

### Clinical assessment and phenotyping

Clinical phenotyping was conducted for all participants. A senior psychiatrist collected demographic information, current medical history, psychological developmental history, and family psychiatric history through structured interviews. The Mini-International Neuropsychiatric Interview (M.I.N.I.) was used to evaluate current and lifetime DSM-5 psychiatric disorders. On the same day as fNIRS acquisition, participants completed validated self-report scales assessing depressive symptoms (PHQ-9), anxiety symptoms (GAD-7), and somatic symptom burden (PHQ-15). These scores were also used to compute the Overall Severity of anxiety spectrum (OSOAS) index, computed as a z unit-based composite score summing up the z scores of PHQ-9, GAD-7 and PHQ-15. All assessments were administered independently by the same senior psychiatrist on the same day to ensure consistency.

### fNIRS data acquisition

fNIRS was employed as a non-invasive neuroimaging tool to quantify cortical hemodynamics during the cognitive task^[35]^. To minimize environmental and motion artifacts, data acquisition was conducted in a sound-attenuated room, and participants were instructed to remain relaxed while avoiding subtle head and facial movements. The signals were acquired using a 53-channel continuous-wave system (BS-5000L, Wuhan Zilian Hongkang Co., Ltd., China), with the optode array widely covering the frontal, parietal, and temporoparietal cortices. This source-detector array consisted of 16 dual-wavelength near-infrared sources (690 nm and 830 nm) and 16 detectors arranged in a high-density grid. The optode array was positioned symmetrically over the bilateral frontotemporal scalp according to the international 10–20 system, with the central optode (9) aligned to the FPz reference point. A 3D electromagnetic digitizer (NirMap, Wuhan Zilian Hongkang Co., Ltd., China) was utilized to record the spatial coordinates of key cranial landmarks (Nz, Cz, AL, RL) and all individual optodes. The 3D coordinates of each channel were projected onto the cortical surface of the Montreal Neurological Institute (MNI) standard brain using the NIRS-SPM probabilistic spatial registration framework and mapped to corresponding Brodmann areas. Based on this registration, the 53 channels were grouped into five bilateral regions of interest (ROIs): supplementary motor area (SMA), Broca’s area, dorsolateral prefrontal cortex (DLPFC), frontal eye fields (FEF), and frontopolar area (FPA). Channel-to-ROI mappings are provided in Electronic Supplementary File (ESF), Table 1.

**Table 1.** Demographic and clinical characteristics of patients with anxiety spectrum disorders (ANSD) and healthy controls (HC)\.

| Variables | HC (n = 62) | ANSD (n=96) | $\chi^2$ | df | p-value |
| --- | --- | --- | --- | --- | --- |
| Age (Years) | 38.8 ± 17.2 | 40.0± 12.0 | MWUT | - | 0.093 |
| Male/Female | 24 / 38 | 39/57 | 0.058 | 1/157 | 0.810 |
| single/married | 13 / 49 | 25/71 | 0.531 | 1/157 | 0.470 |
| Education (years) | 12.95 ± 2.00 | 13.31±3.39 | MWUT | - | 0.130 |
| Body mass index (BMI) (kg/m2) | 22.98 ± 2.53 | 22.59±3.58 | MWUT | - | 0.270 |
| GAD-7 | 1.05 ± 0.97 | 11.72±4.08 | MWUT | - | <0.001 |
| PHQ-9 | 1.08 ± 1.03 | 13.20±5.87 | MWUT | - | <0.001 |
| PHQ-15 | 0.27 ± 0.48 | 12.31±5.98 | MWUT | - | <0.001 |

Note: Data are presented as Mean ± Standard Deviation (SD) for continuous variables and as exact counts for categorical variables. Group comparisons were performed using the Mann-Whitney U test (MWUT) for continuous variables and the Pearson Chi-square test for categorical variables.
BMI, Body Mass Index; GAD-7, Generalized Anxiety Disorder-7; PHQ-9, Patient Health Questionnaire-9; PHQ-15, Patient Health Questionnaire-15.

### Data preprocessing pipeline

fNIRS data were processed using NirMaster data analysis software, version V1.1 (Wuhan Yiruide Medical Equipment New Technology Co., Ltd., Wuhan, China) and comprised the following sequential steps^[36]^: (1) Downsampling: The raw signals were downsampled to 10 Hz to eliminate high-frequency electronic noise. (2) Quality Control: Poor-quality channels with a Coefficient of Variation (CV) > 20% were automatically rejected. Participants with more than 20% rejected channels were excluded from subsequent analyses. (3) Artifact Correction and Filtering: Following the conversion of raw optical intensity signals to optical density (OD), spline interpolation was employed to identify and correct transient motion artifacts induced by head movements. Subsequently, a band-pass filter (0.01 - 0.2 Hz) was applied to effectively eliminate physiological noise arising from cardiac pulsations, respiration, and very low-frequency baseline drifts. (4) Concentration conversion: Based on the Modified Beer-Lambert Law (MBLL), the changes in OD were converted into dynamic concentration change time series for HbO, HbR, and HbT^[17]^. (5) Block Averaging and baseline correction: Utilizing the block averaging method, data epochs encompassing the 10-s pre-task baseline period, the active task period, and the post-task recovery period were extracted. A linear baseline trend was estimated by fitting the data segments from 10-s pre-task and 5-s post-task segments to correct for baseline drift. The mean baseline value of the 10-s pre-task period was then subtracted, followed by block averaging. Finally, a 5-second moving average window was applied to smooth the signals. Although the main ROI-level predictive analyses emphasized HbO, HbR and HbT features were also retained for extended sensitivity analyses and machine-learning models to provide a comprehensive evaluation of neurovascular coupling dynamics. In addition, we assessed five activation metrics for HbO, HbR and HbT, i.e., peak (maximum activation amplitude), mean (average activation), integral (total output over time), slope (speed of activation), and temporal variability (Difference). The biological conclusion was similar regardless of whether mean, peak, or integral metrics were used. Peak HbO was prioritized because it reflects maximal task-evoked neurovascular recruitment and showed the strongest group discrimination. Since mean, peak, and integral HbO were highly correlated and yielded concordant effects, the peak-based findings are unlikely to be metric-specific and instead indicate a robust reduction in cortical activation capacity. In contrast to the highly correlated amplitude-based measures (mean, peak, and integral HbO), the slope parameter showed weak or absent associations with these indices and was not significantly related to ANSD. The Difference metric was the most informative fNIRS feature in the Results analyses.

To ensure the absolute high fidelity of the extracted microvascular hemodynamic features and to strictly minimize the influence of motion or instrumental artifacts, an extremely stringent data quality control (QC) protocol was systematically implemented during preprocessing. According to standardized fNIRS guidelines, channels with a coefficient of variation (CV) greater than 20% were automatically marked as low-quality channels; in addition, any participant with more than 20% bad channels across the 53-channel array was completely excluded from all subsequent statistical and pattern-recognition analyses. After this rigorous signal-purity screening, 5 HC and 105 patients within the anxiety spectrum were excluded because of insufficient data quality. Therefore, the final analytical sample used for multivariate robust estimation and machine-learning classification comprised 158 participants, including 62 high-quality HC and 96 quality-verified inpatients within the anxiety spectrum, with the following subgroup distribution: 28 in the GAD group, 34 in the CO group, and 34 in the AD group.

## Statistical analyses

### Analysis of demographic and clinical variables

IBM SPSS Statistics for Windows, Version 26.0 (Armonk, NY: IBM Corp) was utilized for all clinical and demographic statistical computations. The omnibus significance level was set at ɑ = 0.05 (two-tailed). The distributional assumptions of continuous variables were first evaluated using the Shapiro-Wilk normality test. Given that age, years of education, and all clinical scale scores exhibited non-normal distributions across groups, non-parametric statistics were employed for univariate inter-group comparisons. Specifically, the Chi-square (χ2) test was used for categorical data, and the Kruskal-Wallis H test was applied for continuous non-parametric variables. To mitigate the potentially confounding effects of non-normal distributions and extreme outliers on parametric estimates in subsequent regression modeling, continuous fNIRS hemodynamic indicators and clinical scale scores were subjected where needed to transformations, including logarithmic, square root or fractional rank transformation.

### Multivariate analysis of variance and sensitivity analyses

To examine omnibus differences in regional cerebral hemodynamics between ANSD and HC, a covariate-adjusted multivariate analysis framework was used. Diagnostic group was entered as the independent variable, and fNIRS-derived metrics (Difference and Peak) from ten bilateral ROIs were entered as dependent variables. Sex, age, and years of education were included as covariates. Pillai’s Trace was selected because of its robustness to violations of multivariate assumptions. When omnibus effects were significant, follow-up univariate analyses and post-hoc comparisons were performed to identify ROI-specific alterations, with multiple comparisons controlled using FDR correction, where indicated.

To ascertain the robustness of the neuroimaging findings and explicitly exclude the confounding effects of psychotropic medications, sensitivity analyses were performed. The binary use status (yes/no) of five distinct psychopharmacological classes (SSRIs, SNRIs, benzodiazepines, atypical antipsychotics, and mood stabilizers) was incorporated as additional covariates into the baseline multivariate GLM model^[37,38]^. We subsequently evaluated whether the primary diagnostic group effect remained statistically significant after adjusting for these pharmacological covariates.

For the activation channels with significant main effects in the GLM analysis, post-hoc tests were conducted. To test group differences adjusted for covariates, we first fitted a linear model with covariates only (age and sex) on the pooled pair data and extracted the model residuals; all subsequent tests were performed on these residuals. For the residuals’ distribution, variance equality was assessed by Levene’s test. If both groups’ residuals were approximately normal and variances were equal, a two-sample t-test (equal variances) was used; if normal but variances unequal, Welch’s t-test was used; otherwise, the Wilcoxon rank-sum test was applied. To investigate the dimensional associations between clinical symptom severity and regional hemodynamic signal fluctuations, two-tailed Pearson correlation coefficients were used. Furthermore, to explore the impact of regional hemodynamic fluctuations on specific clinical symptom domains, Multiple Linear Stepwise Regression analyses were conducted with the standardized Difference and Peak indicators across all ROIs as candidate independent variables. The entry and removal criteria for the stepwise feature selection were set at p < 0.05 and p > 0.10, respectively. Collinearity was checked using the Variance Inflation Factor (VIF), with a stringent threshold of VIF < 2.0^[39]^. Binary logistic regression with automatic forward stepwise likelihood ratio selection was executed to determine the most important predictors of ANSD. The discriminative performance of the model was evaluated utilizing Receiver Operating Characteristic (ROC) and Precision-Recall (PR) curves^[40]^.

Subsequently, machine learning classification algorithms, including Random Forest (RF), Support Vector Machine (SVM), and Partial Least Squares discriminant analysis (PLS-DA) were implemented and evaluated on independent test subsets to assess model generalizability. The RF model was constructed utilizing maximal 1000 decision trees, and variable importance was extracted to quantify the relative contributions of specific features to the classification accuracy. Before model fitting, the dataset was randomly divided into training and test subsets using an approximately 70:30 split, and model performance was evaluated on the held-out test subset. The Support Vector Machine (SVM) classifier was implemented using a Radial Basis Function (RBF) kernel with the penalty parameter (C) set to 4.0 and gamma (γ) set to 0.05. Model performance was evaluated using 10-fold cross-validation, in which the dataset was randomly divided into ten subsets, with nine subsets used for training and one for testing in each iteration. Performance metrics were averaged across all folds to assess model generalizability and minimize overfitting.

Furthermore, PLS-DA was performed to extract latent components maximizing inter-group variance (with oversampling of the minority group in the training sample to adjust class imbalance). Variable Importance in Projection (VIP) scores were computed, with a threshold of VIP > 1.0 delineating the most important features driving group separation. The internal validity of the PLS-DA models and the absence of overfitting were verified utilizing permutation tests (by assessing the intercepts of the R2 and Q2 fitting lines)^[41,42]^. All statistical and machine learning algorithms were executed utilizing SPSS (version 30) and STATISTICA 14.0 software.

### GLMs, psychophysiological interaction (PPI), and graph-theoretical network analyses

Individual-level task-evoked activation was estimated using a GLM implemented in the NIRS-KIT toolbox^[43]^. For each channel, the preprocessed time courses of HbO and HbR concentration changes were entered as dependent variables. The design matrix consisted of the following regressors: (1) a task-related regressor generated by convolving the stimulus timing of the VFT with the canonical hemodynamic response function (HRF); and (2) nuisance regressors, including polynomial drift terms (a constant term and a linear term) to account for low-frequency drifts. To address temporal autocorrelation in the residuals, an AR(1) model was applied for pre-whitening. Model parameters (beta values) were estimated using ordinary least squares. For each channel, the contrast of beta estimates between the VFT condition and the baseline condition was computed as the index of task-induced activation magnitude. These contrast values were subsequently submitted to group-level statistical analyses.

We used an Analysis of Covariance (ANCOVA) to control age and gender differences across the HC, CO, GAD, and AD groups. This approach allowed us to treat both variables as covariates and minimize their confounding effects during intergroup analysis. For the activation channels with significant main effects in the ANCOVA analysis, post-hoc tests were conducted. To test group differences adjusted for covariates, we first fitted a linear model with covariates only (age and sex) on the pooled pair data and extracted the model residuals; all subsequent tests were performed on these residuals. For the residuals’ distribution, variance equality was assessed by Levene’s test. If both groups’ residuals were approximately normal and variances were equal, a two-sample t-test (equal variances) was used; if normal but variances unequal, Welch’s t-test was used; otherwise, the Wilcoxon rank-sum test was applied.

Task-modulated functional connectivity was assessed using a generalized PPI (gPPI) analysis^[44]^. To enhance sensitivity to neural activity and to suppress systemic physiological confounds, the difference signal between HbO and HbR concentration changes (HbDiff = HbO − HbR) was used as the hemodynamic measure for the connectivity analysis^[45,46]^. HbDiff was selected because it combines information from both HbO and HbR signals and may provide a more specific representation of task-related oxygenation changes than the HbO signal alone^[31]^. For each participant, the HbDiff time series of all channels were further detrended, band-pass filtered (4th-order Butterworth, 0.01–0.2 Hz), and z-score normalized. The VFT task regressor was constructed as a boxcar function modeling the four 15-s verbal fluency blocks (onset at 30 s), which was subsequently convolved with the canonical hemodynamic response function (SPM spm_hrf) and z-scored to account for the hemodynamic delay. For each seed channel, a PPI regressor was generated as the element-wise product of the seed HbDiff time course and the convolved task regressor. A separate GLM was then fitted for each target channel, with a design matrix consisting of the convolved task term, the seed time series, the PPI interaction term, and a constant. The beta coefficient of the PPI term (β₃) was estimated via ordinary least squares and taken as the index of task-induced connectivity strength between the seed and the target. This procedure was iterated over all channel pairs, yielding an N×N connectivity matrix of raw beta values, with self-connections set to zero. The matrix was symmetrized by averaging with its transpose, and the diagonal was set to zero. The statistical significance of each connection was evaluated using a t-test on the PPI beta estimates. Multiple comparisons across all unique channel pairs were corrected using the FDR procedure (q < 0.05). The resulting thresholded connectivity matrix was retained for subsequent group-level network analyses.

For each participant, a weighted undirected functional brain network was constructed from the FDR-corrected gPPI beta matrix (q < 0.05). Nodes corresponded to fNIRS channels and edges represented significant task-modulated functional connections. Three nodal metrics were computed using GRETNA: weighted clustering coefficient, nodal local efficiency, and nodal efficiency. The weighted clustering coefficient was computed according to the algorithm of Onnela et al.^[47]^, as implemented in GRETNA, to quantify the intensity of local triangular connectivity around each node. Nodal local efficiency was defined as the global efficiency of the subgraph formed by the node’s immediate neighbors, reflecting local information processing capacity and network tolerance. Nodal efficiency was calculated as the average of the inverse shortest path lengths from a given node to all other nodes, characterizing the node’s ability to communicate in parallel across the network. For all efficiency measures, edge weights were converted to connection lengths (distances) using a reciprocal transformation: for existing connections. These three nodal metrics were derived for every channel per subject.

Between-group differences in each nodal metric were assessed using independent-sample t-tests. To correct for multiple comparisons across all channels, the resulting p-values were adjusted using the Benjamini–Hochberg false discovery rate (FDR) procedure, with a significance threshold of q < 0.05. Channels that survived FDR correction were reported as showing statistically significant group effects.

## Results

### Demographic and clinical characteristics

Demographic and clinical characteristics are summarized in **Table 1**. The final sample included 158 participants, consisting of 62 HC and 96 patients with ANSD. The two groups did not differ significantly in sex distribution, age, marital status, years of education, or body mass index. Patients with ANSD showed significantly higher GAD-7, PHQ-9, and PHQ-15 scores than HC.

### Group differences in hemodynamic activation patterns

Covariate-adjusted multivariate analyses were used to test overall group differences in hemodynamic features (**Table 2**). After controlling for sex, age, and years of education, the ANSD and HC groups differed significantly in overall HbO Difference and Peak models. HbO and HbT showed significant group differences for HbR Difference, HbT Difference, and HbT Peak, whereas HbR Peak was not significant.

**Table 2.** Results of the multivariate GLM analysis comparing hemodynamic features between anxiety spectrum disorders and healthy controls.

| Hemodynamic Indicators | F | df | p |
| --- | --- | --- | --- |
| Difference HbO | 6.35 | 10/144 | <0.001 |
| Peak HbO | 3.18 | 10/144 | 0.001 |
| Difference HbR | 5.48 | 10/144 | <0.001 |
| Peak HbR | 1.20 | 10/144 | 0.294 |
| Difference HbT | 5.59 | 10/144 | <0.001 |
| Peak HbT | 2.99 | 10/144 | 0.002 |

Covariate-effect analyses indicated that age had significant main effects on HbO Difference (p = 0.002), HbR Difference (p < 0.001), and HbT Difference (p = 0.008) models. Sex showed significant main effects in the HbO Peak (p = 0.001) and HbT Peak (p = 0.001) models, whereas years of education was not significant in any model.

Univariate GLM analyses identified region-specific alterations in the core hemodynamic features (**Table 3**). For the Difference metric, patients showed significantly greater signal fluctuation than controls in bilateral DLPFC, right Broca’s area, bilateral SMA, and right FEF. For the Peak metric, patients showed significantly lower peak activation in bilateral DLPFC, bilateral FPA, and bilateral Broca’s area, together with a trend toward increased peak values in the right FEF. ESF Table 2 reports the significant FDR-corrected HbR and HbT results examining ANSD patients versus HC. There were no significant differences in peak HbR responses, while the HbR differences showed significant results in different areas. The HbT peak and difference metrics showed aberrations in ANSD in parallel with the HbO findings.

**Table 3.**
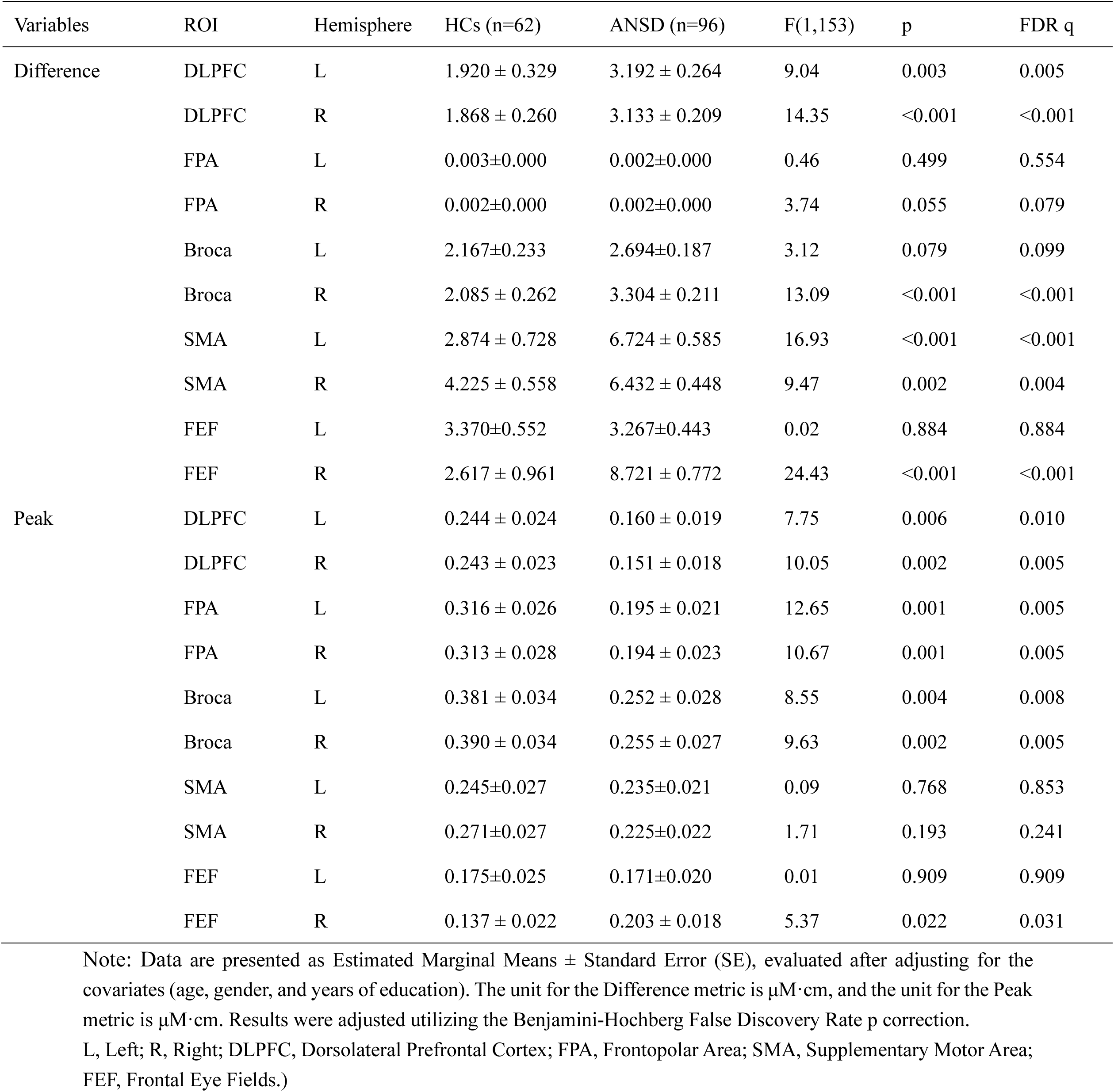
Results of Univariate GLM analyses delineating region-specific hemodynamic alterations in HbO between anxiety spectrum disorders (ANSD) and healthy controls (HC).

| Variables | ROI | Hemisphere | HCs (n=62) | ANSD (n=96) | F(1,153) | p | FDR q |
| --- | --- | --- | --- | --- | --- | --- | --- |
| Difference | DLPFC | L | 1.920 ± 0.329 | 3.192 ± 0.264 | 9.04 | 0.003 | 0.005 |
|  | DLPFC | R | 1.868 ± 0.260 | 3.133 ± 0.209 | 14.35 | <0.001 | <0.001 |
|  | FPA | L | 0.003±0.000 | 0.002±0.000 | 0.46 | 0.499 | 0.554 |
|  | FPA | R | 0.002±0.000 | 0.002±0.000 | 3.74 | 0.055 | 0.079 |
|  | Broca | L | 2.167±0.233 | 2.694±0.187 | 3.12 | 0.079 | 0.099 |
|  | Broca | R | 2.085 ± 0.262 | 3.304 ± 0.211 | 13.09 | <0.001 | <0.001 |
|  | SMA | L | 2.874 ± 0.728 | 6.724 ± 0.585 | 16.93 | <0.001 | <0.001 |
|  | SMA | R | 4.225 ± 0.558 | 6.432 ± 0.448 | 9.47 | 0.002 | 0.004 |
|  | FEF | L | 3.370±0.552 | 3.267±0.443 | 0.02 | 0.884 | 0.884 |
|  | FEF | R | 2.617 ± 0.961 | 8.721 ± 0.772 | 24.43 | <0.001 | <0.001 |
| Peak | DLPFC | L | 0.244 ± 0.024 | 0.160 ± 0.019 | 7.75 | 0.006 | 0.010 |
|  | DLPFC | R | 0.243 ± 0.023 | 0.151 ± 0.018 | 10.05 | 0.002 | 0.005 |
|  | FPA | L | 0.316 ± 0.026 | 0.195 ± 0.021 | 12.65 | 0.001 | 0.005 |
|  | FPA | R | 0.313 ± 0.028 | 0.194 ± 0.023 | 10.67 | 0.001 | 0.005 |
|  | Broca | L | 0.381 ± 0.034 | 0.252 ± 0.028 | 8.55 | 0.004 | 0.008 |
|  | Broca | R | 0.390 ± 0.034 | 0.255 ± 0.027 | 9.63 | 0.002 | 0.005 |
|  | SMA | L | 0.245±0.027 | 0.235±0.021 | 0.09 | 0.768 | 0.853 |
|  | SMA | R | 0.271±0.027 | 0.225±0.022 | 1.71 | 0.193 | 0.241 |
|  | FEF | L | 0.175±0.025 | 0.171±0.020 | 0.01 | 0.909 | 0.909 |
|  | FEF | R | 0.137 ± 0.022 | 0.203 ± 0.018 | 5.37 | 0.022 | 0.031 |

Note: Data are presented as Estimated Marginal Means ± Standard Error (SE), evaluated after adjusting for the covariates (age, gender, and years of education). The unit for the Difference metric is $\mu\text{M}\cdot\text{cm}$ , and the unit for the Peak metric is $\mu\text{M}\cdot\text{cm}$ . Results were adjusted utilizing the Benjamini-Hochberg False Discovery Rate p correction.
L, Left; R, Right; DLPFC, Dorsolateral Prefrontal Cortex; FPA, Frontopolar Area; SMA, Supplementary Motor Area; FEF, Frontal Eye Fields.)

Sensitivity analyses were performed to evaluate whether psychotropic medication use confounded the primary findings. Among the 96 patients with ANSD, 76 (79.2%) used escitalopram (n = 51), sertraline (n = 12), paroxetine (n = 11), venlafaxine (n = 19), duloxetine (n = 10), 83 (86.5%) used benzodiazepines, 48 (50.0%) used atypical antipsychotics, and 6 (6.3%) used mood stabilizers. Five medication classes (selective 5-HT reuptake inhibitors, selective noradrenaline inhibitors, benzodiazepines, atypical antipsychotics, and mood stabilizers) were added as binary covariates to the baseline multivariate models None of the medication covariates showed significant main or univariate effects on the activation features even without FDR p correction (all p > 0.05). After simultaneous adjustment for demographic and medication covariates, the group effects remained significant for HbO Difference (Pillai’s Trace = 0.306, F(10,144) = 6.346, p < 0.001) and HbO Peak (Pillai’s Trace = 0.181, F(10,144) = 3.181, p = 0.001). Among the three ANSD subgroups, FDR-significant omnibus effects were confined to the difference metrics in HbO, HbR, and HbT (ESF Table 3); FDR-corrected post-hoc comparisons showed that the AD group differed from GAD and CO, whereas no GAD-versus-CO comparison survived. These changes were further explored using gPPI and graph theory analyses (see below).

### Classification of diagnosis ANSD versus controls

Table 4 shows the results of logistic binary regression analysis predicting ANSD (with controls as the reference group). To identify the most important independent fNIRS biomarkers for ANSD, a forward stepwise binary logistic regression model was constructed. The six significant predictors were Difference_FEF_R, Difference_Broca_R, and Peak_FEF_L (positively associated with ANSD) and Difference_FEF_L, Difference_FPA_R, and Peak_Broca_R (negatively associated). This model yielded a Nagelkerke R2 of 0.607 and an AUC ROC of 0.911 (95% CI: 0.865-0.958, p < 0.001; ESF, Figure 1 A) with overall accuracy of 84.8%, 81.3% sensitivity, and 93.5% specificity. The precision-recall curve (ESF Figure 1 B) indicated a precision of 95.1% at 81.3% recall.

**Figure 1.**
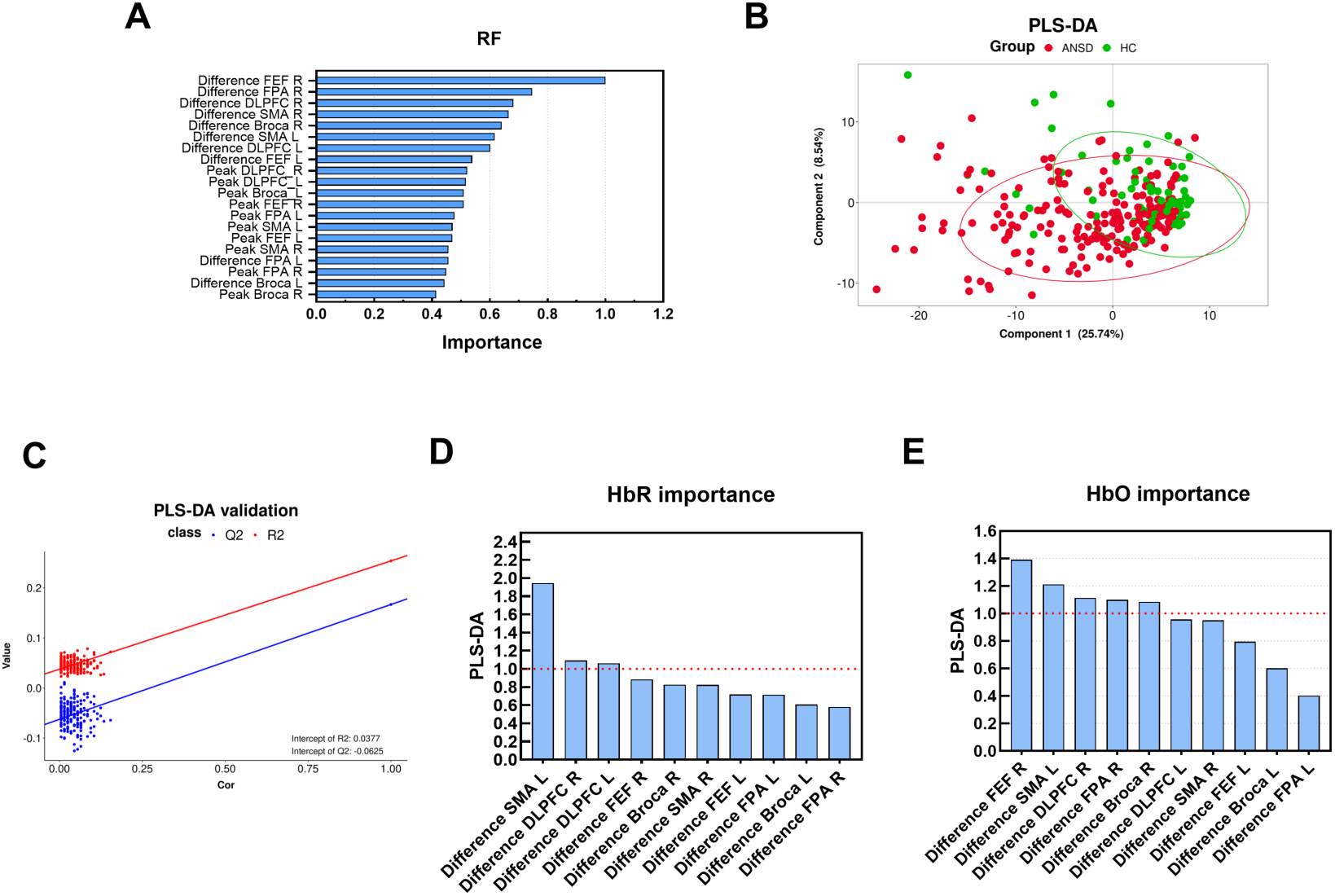
Feature extraction and validation of the machine learning classification models. (1A) Variable importance plot derived from the Random Forest (RF) model, ranking the relative contributions of hemodynamic HbO features. (1B) Variable Importance in Projection (VIP) scores from the Partial Least Squares Discriminant Analysis (PLS-DA) model for HbO features. The red dashed line represents the critical VIP threshold of 1.0. (1C) VIP scores extracted from the PLS-DA model specifically constructed for HbR features (1D) Two-dimensional PLS-DA score plot displaying the spatial distribution and separation between participants with anxiety spectrum disorders (ANSD, red dots) and healthy controls (HC, green dots) along the first two principal components. (1E) Permutation test results validating the PLS-DA model. The positive slope of the R2 line and the negative intercept of the Q2 line (Q2 intercept = -0.0665) indicate the absence of model overfitting.

**Table 4.** Results of binary logistic regression analysis with anxiety spectrum disorder as the outcome and controls as reference group.

| Predictors | $\beta$ | SE | Wald<br>(df=1) | p | Exp(B) | 95% C.I. for Exp(B) | |
| --- | --- | --- | --- | --- | --- | --- | --- |
|  |  |  |  |  |  | Lower | Upper |
| Difference FEF R | 1.842 | 0.371 | 24.648 | <0.001 | 6.307 | 3.048 | 13.049 |
| Difference Broca R | 1.035 | 0.313 | 10.890 | 0.001 | 2.814 | 1.522 | 5.202 |
| Difference _FPA_R | -0.854 | 0.277 | 9.511 | 0.002 | 0.426 | 0.247 | 0.733 |
| Difference FEF L | -1.514 | 0.363 | 17.410 | <0.001 | 0.220 | 0.108 | 0.448 |
| Peak FEF L | 0.642 | 0.275 | 5.469 | 0.019 | 1.900 | 1.110 | 3.254 |
| Peak Broca R | -0.760 | 0.273 | 7.717 | 0.005 | 0.468 | 0.274 | 0.800 |

Note: $\beta$ , regression coefficient; SE, standard error; Wald, Wald $\chi^2$ statistic; Exp(B), odds ratio; CI, confidence interval; FEF, frontal eye fields; FPA, frontopolar area; L, left; R, right. Controls served as the reference group.

Figure 1 shows the results of random Forest and PLS-DA analysis performed on the combined HbO Peak and Difference features or on the latter alone. Figure 1A shows the importance plot obtained using random forest. The random forest classifier (1000 trees, maximum tree size = 100) was used to discriminate HC from ANSD cases using the fNIRS HbO features and clinical variables. The model converged rapidly, with the training and test misclassification rates stabilizing after approximately 250 trees. Across all subjects, the model correctly classified 48 of 62 HC (77.4%) and 86 of 96 affective disorder cases (89.6%). The overall classification accuracy for the complete sample was 84.8% (134/158). These findings indicate that the combined fNIRS feature set provides good discrimination between ANSD and controls. The five most influential features were Difference_FEF_R (normalized importance = 100%), Difference_FPA_R, Difference_DLPFC_R, Difference_SMA_R, and Difference_Broca_R. The radial-basis-function SVM model correctly classified controls and patients with an overall test accuracy of 77.8%.

Figure 1B, 1D and 1E show the results of PLS-DA analyses using the HbO Difference only (Figure 1). The PLS-DA score plot (Panel DB) showed a partial separation between ANSD and controls. Although overlap between groups was observed, the distribution of cases indicated a clear tendency toward class discrimination. The PLS model extracted two significant latent components, which together explained 36.98% of the variance in the outcome variable (R²Ycum = 0.3698) and 42.87% of the variance in the predictors (R²Xcum = 0.4287). The model showed modest predictive performance (Q²cum = 0.1930). The permutation test (Panel 1C) demonstrated adequate model validity, with a negative Q² intercept and a low R² intercept, indicating that the observed classification was unlikely due to overfitting. Five variables exceeded the VIP > 1.0 threshold (Figure 1D): Difference_FEF_R, Difference_SMA_L, Difference_DLPFC_R, Difference_FPA_R, and Difference_Broca_R. Interestingly, also the HbR Difference showed some discriminatory validity (Figure 1D). Two significant latent variables were extracted (cumulative R2Y = 0.307; cumulative Q2 = 0.229), with VIP > 1.0 for Difference_SMA_L (VIP = 1.947), Difference_DLPFC_R (VIP = 1.091), and Difference_DLPFC_L (VIP = 1.062).

ESF, Figure 2 shows individualized feature-contribution profiles for four representative participants, including one HC and three patients. HC showed negative or near-baseline contributions across the HbO Difference metrics. In contrast, patients showed heterogeneous contribution patterns: patient 1 was primarily driven by a high positive contribution from Difference_FEF_R; patient 2 showed dominant contribution from Difference_SMA_L; and patient 3 showed a combined pattern driven by both Difference_FEF_R and Difference_SMA_L.

**Figure 2.**
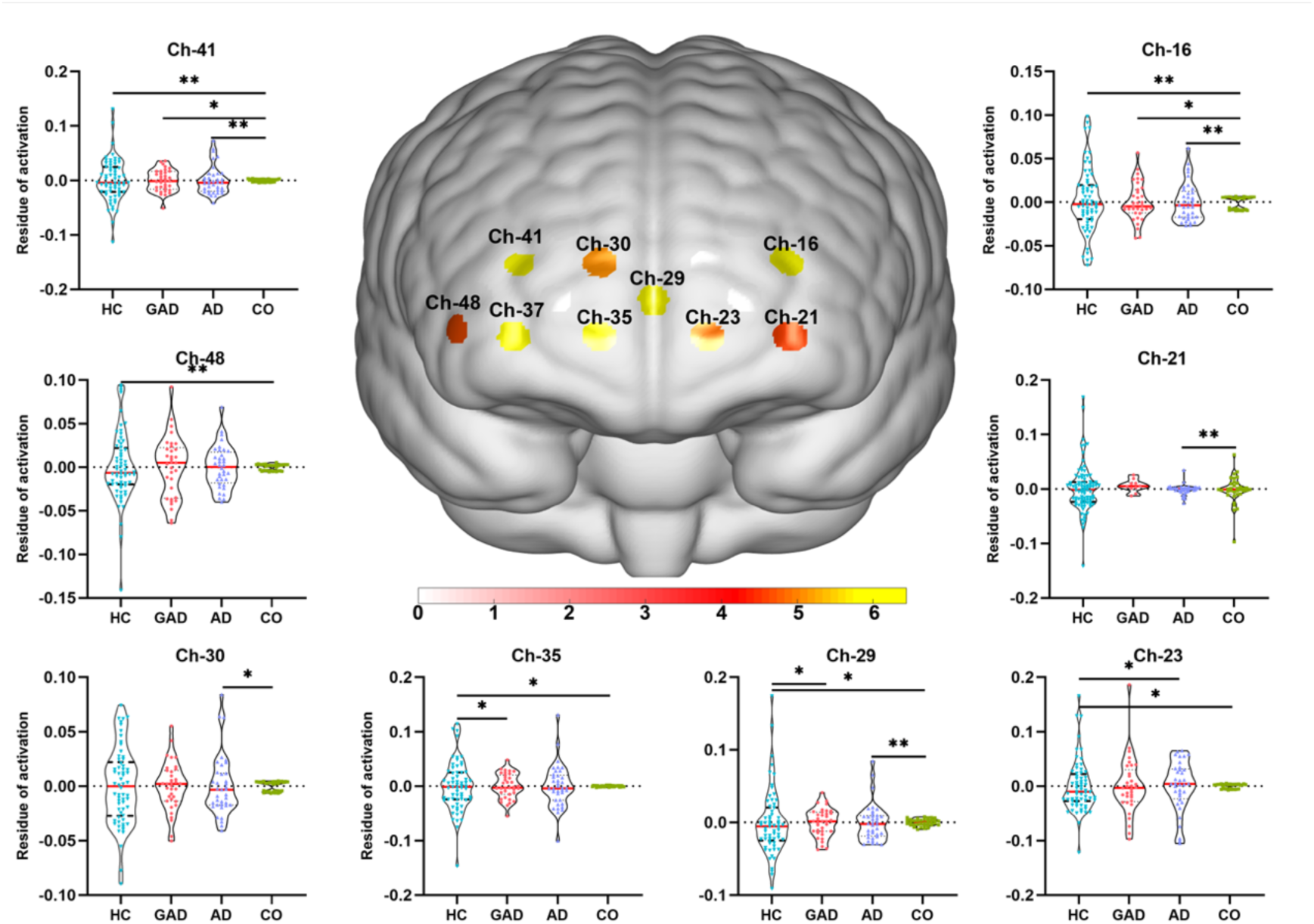
Brain regions showing significant group differences in task-evoked cortical activation across the anxiety spectrum. This figure presents the brain activation map and corresponding post-hoc comparisons derived from the analysis of covariance. The brain map displays the F-value distribution of channels that remained significant after false discovery rate correction, highlighting regions with significant group differences. The eight violin plots show the post-hoc pairwise comparisons for each significant channel, showing the activation values across healthy controls (HC), generalized anxiety disorder (GAD), anxiety–depression comorbidity (CO), and anxious depression (AD) subgroups.

### Results of multiple regression analyses with symptom domain scores as dependent variables

Table 5 examines the associations between regional features and clinical symptom severity. Multiple stepwise linear regression models were constructed for GAD-7, PHQ-9, PHQ-15, and OSOAS. We found that 35.2% of the variance in GAD-7 was explained by Difference_FEF_R and Difference_SMA_L (positively), Difference_FPA_R, Difference_FEF_L, and Difference_DLPFC_L (negatively). We found that 39.3% of the variance in PHQ-9 was explained by Difference_FEF_R, Difference_DLPFC_R, and Difference_SMA_L (positively), Peak_DLPFC_R and Difference_FEF_L (negatively). A large part of the variance in the PHQ-15 was associated with Difference_FEF_R, Difference_SMA_L, and Difference_DLPFC_R (positively), Peak_DLPFC_R and Difference_FEF_L (inversely). Difference_FEF_R and Difference_SMA_L (positively), Difference_FEF_L and Peak_DLPFC_R (negatively) explained 37.6% of the variance in OSOAS.

**Table 5.** Stepwise multiple linear regression models predicting clinical symptom severity using regional signal fluctuations as explanatory variables.

| Dependent Variables | Explanatory Variables | Coefficient statistics |  |  | Model statistics |  |  |  |
| --- | --- | --- | --- | --- | --- | --- | --- | --- |
| | | $\beta$ | t | p | R<sup>2</sup> | F | df | p |
| #1. GAD-7 | Model |  |  |  |  |  |  |  |
|  | Difference_FEF_R | 0.384 | 5.167 | <0.001 | 0.352 | 16.506 | 5/152 | < 0.001 |
|  | Difference_SMA_L | 0.264 | 3.722 | <0.001 |  |  |  |  |
|  | Difference_FPA_R | -0.202 | -2.884 | 0.005 |  |  |  |  |
|  | Difference_FEF_L | -0.197 | -2.706 | 0.008 |  |  |  |  |
|  | Difference_DLPFC_L | -0.195 | -2.778 | 0.006 |  |  |  |  |
| #2. PHQ-9 | Model |  |  |  |  |  |  |  |
|  | Difference_FEF_R | 0.384 | 5.260 | <0.001 | 0.393 | 19.675 | 5/152 | < 0.001 |
|  | Peak DLPFC R | -0.245 | -3.693 | <0.001 |  |  |  |  |
|  | Difference DLPFC R | 0.241 | 3.513 | 0.001 |  |  |  |  |
|  | Difference_SMA_L | 0.240 | 3.536 | 0.001 |  |  |  |  |
|  | Difference_FEF_L | -0.210 | -3.034 | 0.003 |  |  |  |  |
| #3. PHQ-15 | Model |  |  |  |  |  |  |  |
|  | Difference_FEF_R | 0.374 | 4.974 | < 0.001 | 0.353 | 16.579 | 5/152 | <0.001 |
|  | Peak DLPFC R | -0.291 | -4.253 | < 0.001 |  |  |  |  |
|  | Difference_FEF_L | -0.284 | -3.965 | < 0.001 |  |  |  |  |
|  | Difference_SMA_L | 0.193 | 2.747 | 0.007 |  |  |  |  |
|  | Difference_DLPFC_R | 0.166 | 2.339 | 0.021 |  |  |  |  |
| #4. OSOAS | Model |  |  |  |  |  |  |  |
|  | Difference_FEF_R | 0.472 | 6.701 | < 0.001 | 0.376 | 23.090 | 4/153 | < 0.001 |
|  | Difference FEF L | -0.276 | -3.950 | < 0.001 |  |  |  |  |
|  | Difference SMA L | 0.251 | 3.659 | < 0.001 |  |  |  |  |
|  | Peak DLPFC R | -0.208 | -3.225 | 0.002 |  |  |  |  |

Note: GAD-7, Generalized Anxiety Disorder-7; PHQ-9, Patient Health Questionnaire-9; PHQ-15, Patient Health Questionnaire-15; OSOAS, Overall Severity of the Anxiety Spectrum; DLPFC, dorsolateral prefrontal cortex; FEF, frontal eye fields; FPA, frontopolar area; SMA, supplementary motor area; L, left; R, right; $\beta$ , standardized regression coefficient; t, t statistic; R<sup>2</sup>, coefficient of determination; F, model F statistic; df, degrees of freedom. GAD-7, PHQ-9, and PHQ-15 were Z-standardized, whereas OSOAS (overall severity of anxiety spectrum) was modeled using raw scores. “Difference” predictors represent Z-scores derived from fractional ranks, whereas “Peak” predictors represent direct Z-scores.

### Task-evoked cortical activation maps across ANSD

In contrast to the above analyses on HbO/HbR Peak and Differences, the GLM-derived activation maps, functional connectivity, and network topology showed not only differences between controls and ANSD but also some important differences among the three subgroups. Therefore, these results are displayed in controls and the three ANSD subgroups. As shown in ESF, Figure 3, the control group demonstrated widespread and robust positive activation across almost all channels. Conversely, the ANSD group showed reduced positive activation compared to the control group, with prominent negative activation observed in the right prefrontal area. The CO group displayed the most distinct pattern, characterized by strong positive activation only in few channels (19, 25, and 40), while the majority of the remaining channels exhibited deactivation or no significant activation.

**Figure 3.**
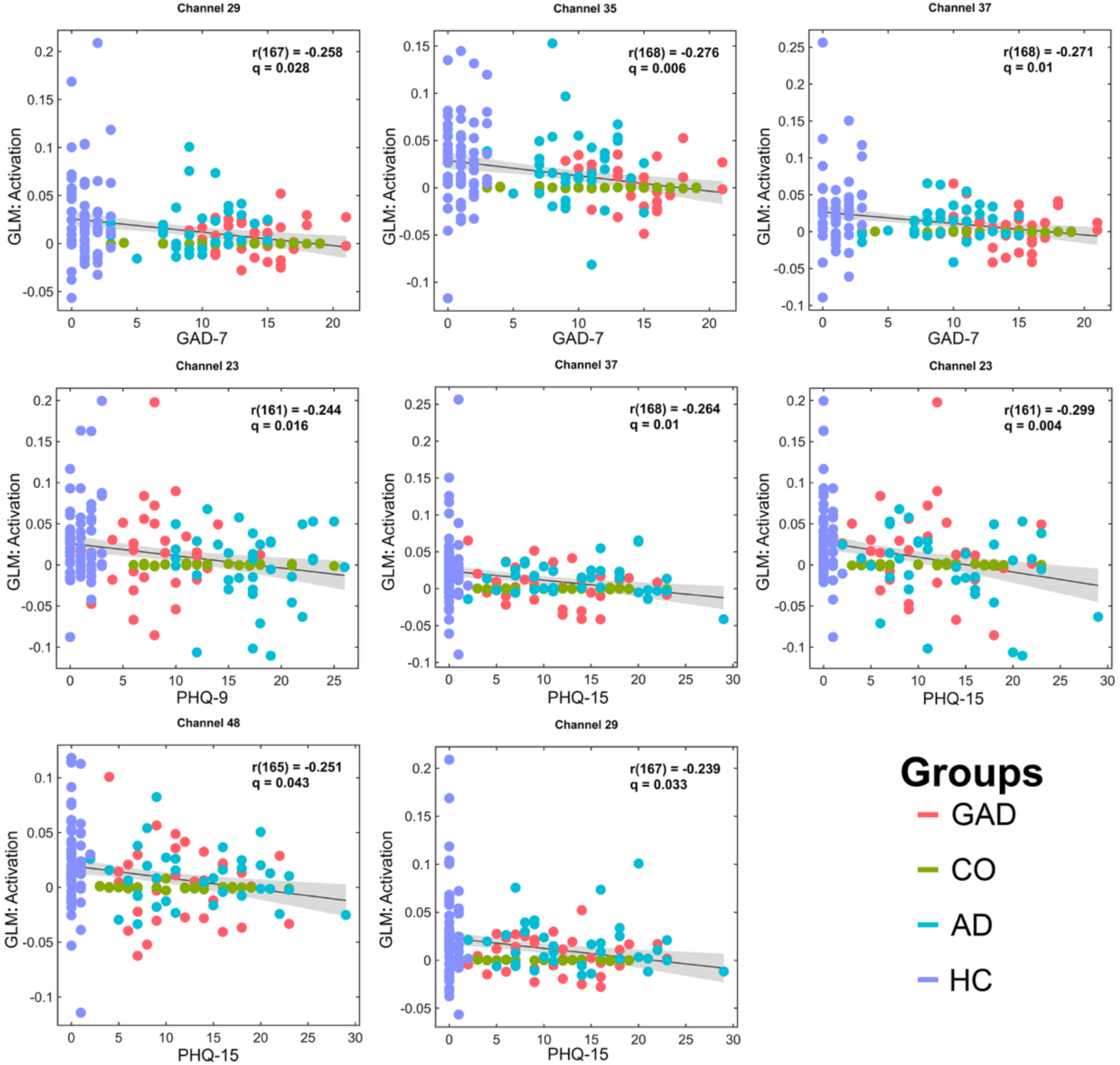
Associations between regional GLM Activation (β₁) and clinical symptom severity across the anxiety Spectrum . Correlations between β value and clinical questionnaires. All results were p corrected for false Discovery Rate (q values). HC: healthy controls, GAD: generalized anxiety disorder, CO: anxiety–depression comorbidity, AD: anxious depression

These findings were further validated by ANCOVA and post-hoc pairwise comparisons (Figure 2). ANCOVA revealed significant main effects of groups across nine prefrontal channels (16, 21, 23, 29, 30, 35, 37, 41, and 48). In the post-hoc comparisons, the CO group demonstrated a distinct activation profile significantly different from the other three groups, generally exhibiting the lowest residual activation levels. Specifically, the GAD group showed significantly lower activation compared to the control group in channels 29, 35, and 41. The AD group exhibited a broader range of reduced activation relative to the control group, spanning not only channel 23 but also channels 29, 30, 35, and 48.

Consequently, we performed Pearson correlation analyses between the extracted β1-values and clinical scale scores. All results were corrected for multiple comparisons using FDR (Figure 3). Negative correlations were observed between prefrontal activation and anxiety, depressive, and physiosomatic rating scales. Specifically, GAD-7 scores were significantly negatively correlated with activation levels in channels 29, 35, and 37 (r ranging from -0.258 to -0.276, all q < 0.05). PHQ-9 scores were negatively correlated with activation in channels 23 and 37, whilst PHQ-15 scores were negatively correlated with activation in channels 23, 29, and 48 (r ranging from -0.239 to -0.299, all q < 0.05). Notably, channel 23 consistently exhibited significant negative correlations with both PHQ-9 and PHQ-15 scores (q values as low as 0.016 and 0.004). In summary, reduced activation in prefrontal cortical regions, particularly channels 23, 29, 35, 37, and 48, is significantly correlated with more severe anxiety, depression, and physiosomatic symptoms.

### Task-based functional connectivity networks using gPPI analysis

To further explore the network-level mechanisms underlying the differences in regional activation observed previously, we constructed task-based functional connectivity networks using gPPI analysis and conducted inter-group comparisons (Figures 4 **and 5**). The control group displayed connectivity strengths within a relatively narrow range (from -0.1 to 0.1), whereas the CO and GAD groups demonstrated both stronger positive and negative functional connections (with color bar spans reaching 0.5 and 0.3, respectively).

**Figure 4.**
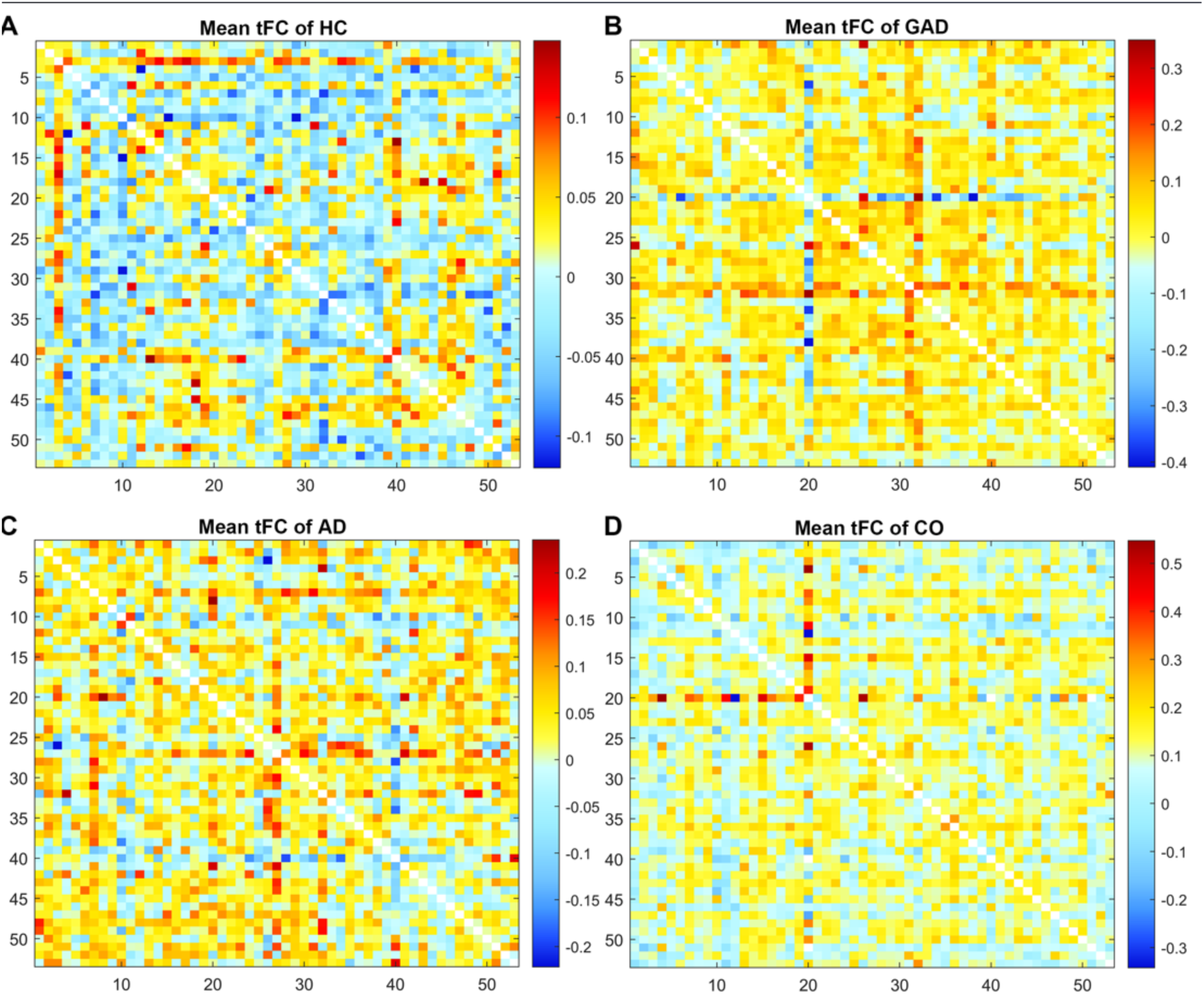
Group differences in task-based functional connectivity (tFC) across healthy controls and anxiety-spectrum Disorders. Panels A–D display the group-averaged task-based functional connectivity matrices for healthy controls (HC), anxiety–depression comorbidity (CO), generalized anxiety disorder (GAD), and anxious depression (AD), respectively. Compared with HC, all anxiety-spectrum groups show more widespread positive interregional connectivity. The strongest and most diffuse connectivity pattern was observed in the CO group.

**Figure 5.**
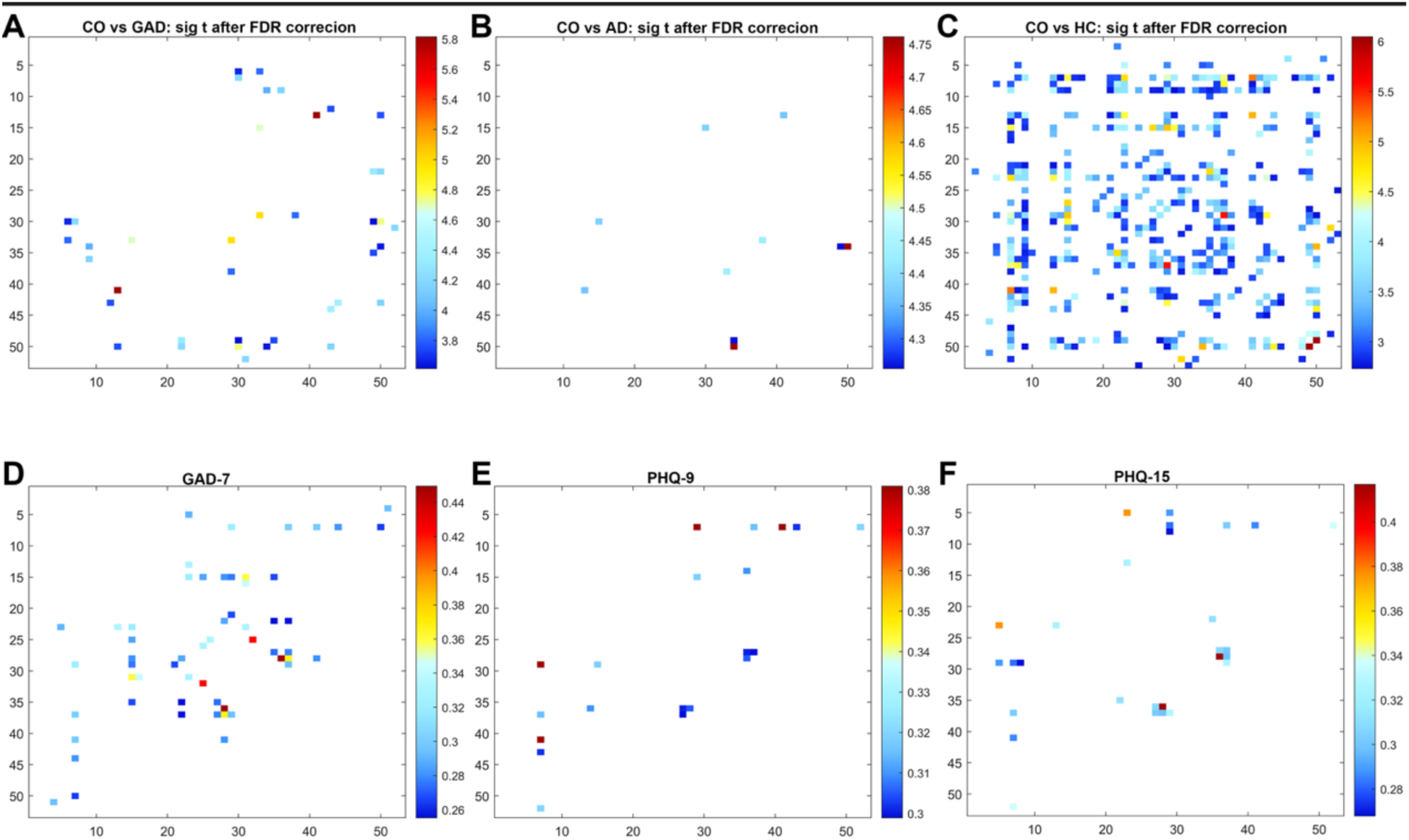
Channel-to-channel functional connectivity differences between anxiety-spectrum groups and significant Associations Between Task-Based Functional Connectivity and Clinical Symptom Severity Across the Anxiety Spectrum. 5A: comorbid anxiety-depression group (CO) vs. generalized anxiety disorder group (GAD); 5B: CO vs. anxious depression (AD) group; 5C: CO vs. healthy controls (HC). The colorbar in the figure represents the t-values from comparisons between different functional connectivity networks. Panels 5D, 5E, 5F display channel-to-channel functional connectivity pairs significantly correlated with anxiety (GAD-7), depressive (PHQ-9), and somatic (PHQ-15) symptom severity, respectively, after FDR correction. The largest number of significant associations was observed for GAD-7, indicating a close relationship between altered functional connectivity and anxiety severity. Significant but less extensive connectivity–symptom associations were also observed for PHQ-9 and PHQ-15, suggesting that network dysregulation contributes to depressive and somatic symptom dimensions across the anxiety spectrum.

Subsequent inter-group statistical comparisons (FDR corrected) revealed clearer patterns of network alterations (Figure 5A, 5B, 5C). The most extensive and significant differences in functional connectivity were observed between the CO group and control group, with the significantly altered node pairs distributed diffusely across the prefrontal network and t-values reaching up to 6 (Figure 5C). Comparisons between the CO and GAD groups also revealed a moderate number of significantly different connections (t-values ranging from approximately 3.8 to 5.8), suggesting partial network heterogeneity between these two states. The number of significantly different connections between the CO and AD groups after FDR correction was sparse, with only a few discrete nodes/edges exhibiting significant differences (t-values ranging from approximately 4.3 to 4.75).

### Graph theory metrics

Building upon the observed differences in task-based functional connectivity, we further calculated graph theory metrics to elucidate the topological reorganization of prefrontal networks across groups (Figure 6). All results were corrected for multiple comparisons using the FDR. The control group consistently exhibited the highest levels across various network topological metrics, whereas the patient groups (particularly the AD and CO groups) displayed significant "disintegration" in information processing and network integration at key nodes. Specifically, regarding the clustering coefficient (Figure 6A), the control group demonstrated significant advantages across multiple channels. AD patients showed significantly lower clustering coefficients in channels 15, 20, 26, and 42 compared to the control group. Additionally, the CO group exhibited a significantly reduced clustering coefficient in channel 20 relative to the HC group (P < 0.05), suggesting impaired local connectivity tightness in the comorbid state. For nodal local efficiency (Figure 6B), inter-group differences were more pronounced. The AD group displayed significantly lower local efficiency in channels 20, 26, and 42 compared to the HC group. Similarly, the CO group showed significant reductions in channel 20, indicating severely impaired efficiency in transmitting information to neighboring nodes in both AD and CO groups.

**Figure 6.**
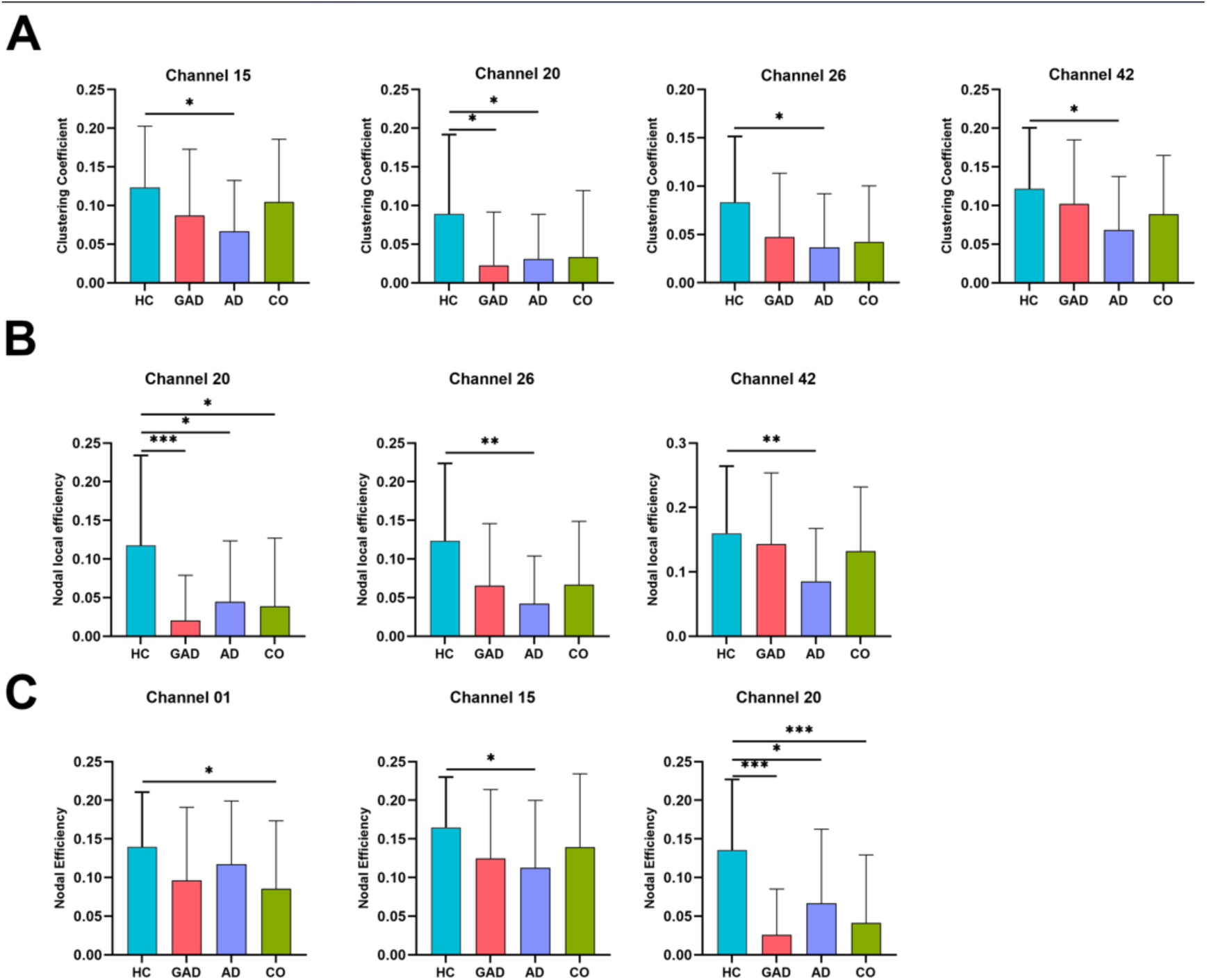
Group comparison of graph theory metrics in task-based functional connectivity networks. 6A: inter-group comparisons of clustering coefficients; 6B: inter-group comparisons of nodal local efficiency; 6C: inter-group comparisons of nodal efficiency. All results are corrected for false discovery rate. *p < 0.05; **p < 0.01; ***p < 0.001

Regarding nodal efficiency, which reflects global information integration capacity (Figure 6C), channel 20 again emerged as a critical hub with the most pronounced differences. The GAD group exhibited a remarkably significant decrease in nodal efficiency at this channel. The CO and AD groups also demonstrated significantly lower nodal efficiency in channel 20 compared to the HC group. Furthermore, the AD group showed significantly lower nodal efficiency in channels 1 and 15, further confirming their disadvantages in global network information transfer.

We further investigated the relationship between tFC strengths and the patients’ clinical symptom domains. We conducted Pearson correlation analyses between edge-level FC values (derived from the gPPI models) and clinical questionnaire scores, with all results corrected using the FDR (Figures 5D, 5E, 5F). The analysis revealed positive correlations between the strength of specific prefrontal node-to-node functional connections and the severity of the three symptom domains. For GAD-7, the correlations were predominantly focused on connections between the bilateral medial prefrontal cortex (mPFC) and the left inferior frontal gyrus (IFG). Notably, channel 23 (MNI: -13, 73, -2; left mPFC/frontal pole), channel 5 (MNI: -51, 30, 33; left IFG, pars triangularis) and channel 13 (MNI: -46, 47, 21) emerged as important hubs. Furthermore, the connection between channel 21 (MNI: -33, 66, -2; left frontal pole) and channel 29 (MNI: 0, 68, 8; midline mPFC) was also significantly associated with anxiety severity. Regarding depressive and physiosomatic symptoms, the correlation network expanded to involve the right prefrontal cortex. PHQ-9 scores showed significant positive correlations with the connection between channel 7 (MNI: -55, 36, 4; left IFG, orbital part) and channel 29 (midline mPFC) (r = 0.381), as well as the connection between channel 28 (MNI: 1, 63, 28; midline mPFC) and channel 36 (MNI: 24, 72, 9; right frontal pole) (r = 0.305). Most notably, PHQ-15 scores exhibited a highly specific correlation pattern focused on the right prefrontal cortex. The strongest correlation was observed between channel 28 and channel 36. Furthermore, the connection involving channel 37 (MNI: 34, 67, -2; right frontal pole) and channel 29 (midline mPFC) was also significantly associated. In summary, the MNI-based spatial mapping confirms that functional connectivity abnormalities within a prefrontal network anchored by the left inferior frontal gyrus (encompassing Brodmann areas 44/45/47) and the bilateral medial frontal/frontopolar regions (encompassing Brodmann areas 10/9/11) constitute the neural substrate for the anxiety, depressive, and physiosomatic symptoms exhibited during verbal fluency task performance.

## Discussion

### Diminished cerebral blood-volume recruitment in ANSD

The first major finding of this study is that ANSD are characterized by lower HbO peak in the DLPFC, Broca, SMA and FEF areas, whilst HbR is unchanged and HbT is reduced. Previous fNIRS studies in anxiety disorders and major depression have consistently reported reduced task-evoked prefrontal HbO responses, particularly during verbal fluency and executive-function paradigms. Herrmann et al. demonstrated bilaterally reduced frontal HbO activation in patients with depression^[48]^. In anxiety disorders, reduced prefrontal hemodynamic responsiveness and altered functional connectivity have likewise been reported, particularly within executive and emotion-regulation networks^[24,49]^. Akiyama et al. observed reduced DLPFC HbO activation in MDD, which was associated with symptom severity and cognitive dysfunction^[50]^. Meta-analytic evidence further confirmed that attenuated HbO responses during cognitive tasks represent one of the most reproducible fNIRS findings in affective and related psychiatric disorders.^23^ While most previous fNIRS studies have focused primarily on task-evoked HbO responses, the present findings extend this literature by demonstrating a concurrent reduction in HbT alongside preserved HbR levels.

HbO reflects oxygenated blood delivery, whereas HbT indexes cerebral blood-volume (CBV) recruitment and vascular expansion. Consequently, the concurrent reductions in HbO and HbT indicate impaired task-evoked perfusion and diminished neurovascular recruitment rather than enhanced oxygen extraction. The absence of significant HbR increases suggests reduced hemodynamic responsiveness and blunted vascular gain during cognitive activation^[51]^.

### Greater temporal hemodynamic variability in ANSD

The second major finding is that ANSD showed a dissociation between reduced HbO Peak responses and increased HbO Difference values, indicating blunted task-evoked hemodynamic recruitment alongside greater temporal variability of the oxygenation response. While previous fNIRS studies in anxiety and depression have reported reduced prefrontal HbO responses during cognitive tasks (see previous section), few studies have examined dynamic fluctuation measures. While reduced peak HbO indicates impaired hemodynamic gain during cognitive activation,^48^ our findings reflect greater temporal variability of the hemodynamic response. This suggests unstable or dysregulated neurovascular dynamics rather than simple hypoactivation and that cortical perfusion responses may be inefficient.

### Reduced task-evoked cortical activation in ANSD

The third major finding is that task-evoked activation, functional connectivity and network topology analyses showed additional aberrations in ANSD, with sometimes significant differences among the three ANSD subgroups. This contrasts with the HbO and HbR assessments discussed above, which showed only minimal differences among the ANSD subgroups. The β1 activation topography shows a widespread and homogeneous positive activation in controls, particularly across the bilateral prefrontal, frontopolar, and dorsolateral prefrontal regions, whereas ANSD is characterized by a significantly lower activation. Hu et al^[24]^. demonstrated abnormal prefrontal activation patterns in GAD, MDD, and their comorbidity. Previous fNIRS studies in depression have reported reduced task-evoked frontal, prefrontal, frontopolar, and dorsolateral prefrontal activation based on GLM-derived activation maps^[48,50,52]^.

Our findings additionally show that the CO group exhibited the lowest and most spatially restricted β₁ activation, indicating the greatest disruption of prefrontal activation and network engagement. Furthermore, ANSD patients showed reduced activation in several prefrontal channels, particularly the left frontal pole (channel 23; MNI: -13, 73, -2; BA10), medial prefrontal cortex (channel 29; MNI: 0, 68, 8; BA10/11), and right frontal pole (channel 35; MNI: 13, 73, -2; BA10). These areas play central roles in self-referential processing, emotion regulation, cognitive flexibility, executive control, and verbal fluency, and substantially overlap with core hubs of the default-mode and executive-control networks^[53–56]^.

### Functional connectivity and network topology in ANSD

Overall, the task-based functional connectivity analyses revealed significantly increased interregional coupling across ANSD compared with controls, indicating enhanced synchronization of cortical oxygenation dynamics during cognitive processing. The comparison of Graph Theory Metrics in Task-Based Functional Connectivity Networks indicated that those patients exhibit a significantly reduced clustering coefficient, nodal local efficiency, and nodal efficiency.

Previous fNIRS studies indicate that anxiety disorders are characterized not only by abnormal regional activation but also by alterations in large-scale cortical connectivity and network organization. For example, Hu et al. demonstrated abnormal prefrontal activation and connectivity patterns in GAD, MDD, and their comorbidity during a verbal fluency task, with the comorbid group showing the most pronounced abnormalities^[24]^. More generally, fNIRS-based network studies have shown that psychiatric disorders are often accompanied by reduced local efficiency, lower clustering coefficients, and disrupted small-world topology, indicating reduced network segregation and impaired information-processing efficiency^[57,58]^.

Interestingly, our fNIRS observations are highly consistent with findings from resting-state and task-based fMRI studies. For example, Sylvester et al. reviewed evidence indicating that anxiety disorders are characterized by dysfunctional large-scale networks involving the salience, executive-control, and default-mode networks^[59]^. Kaiser et al. demonstrated that mood disorders are with increased connectivity together with impaired segregation and integration of cortical systems^[60]^. Graph-theoretical MRI studies in anxiety disorders and major depression have reported reductions in clustering coefficient, local efficiency, and small-world organization, indicating impaired network segregation^[58,61,62]^. These abnormalities have been interpreted as reflecting disrupted large-scale network organization and inefficient communication among distributed cortical systems^[59,60]^.

An important finding of our study is that the comorbidity subgroup showed widespread and pronounced connectivity differences from the control group, involving multiple connections among the left inferior frontal gyrus (IFG; e.g., channel 5, MNI: –51, 30, 33, BA45/47), frontal pole (channel 21, MNI: –33, 66, – 2), and mPFC (channel 29). These regions constitute a network essential for semantic retrieval, inhibitory control, and internal speech generation during verbal fluency^[55,63]^. In the comorbidity group, abnormal strengthening or weakening of these connections may reflect compensatory or maladaptive plasticity.

A particularly important supplementary finding is that our network analyses revealed the AD group showed the most extensive abnormalities, with reduced clustering coefficients in Channels 15, 20, 26, and 42, together with decreased local efficiency and nodal efficiency across multiple nodes. Channel 20 (MNI: -12, 59, 40; left premotor / supplementary motor area, SMA) emerged as a particularly vulnerable hub, exhibiting reduced local and global efficiency in the AD and CO groups. The SMA is involved not only in motor planning and sequence generation for verbal fluency but also works together with the mPFC and IFG as part of a subnetwork for speech execution^[55,64]^. A decreased clustering coefficient at this node implies weakened connectivity among its local neighbors, reflecting that this node’s role as a hub for whole-network information transfer is impaired. Therefore, channel 20 can be considered a critical node in the decentralization of prefrontal networks in ANSD.

Furthermore, these network abnormalities occurred in parallel with reduced HbO and HbT responses, preserved HbR levels, increased HbO/HbR hemodynamic variability, and lower β₁ activation. This combination suggests that ANSD are characterized by maladaptive hyperconnectivity occurring on a background of impaired neurovascular recruitment and unstable regulation. Notably, the CO group exhibited the most severe alterations, showing the lowest activation and the most pronounced network abnormalities, whereas GAD and AD occupied an intermediate position.

### Associations between symptom severity and hemodynamic assessments

A final important finding was that anxiety, depressive, and physiosomatic symptoms were predominantly predicted by the HbO Difference metrics and lower β₁ activation in several prefrontal channels. The strongest and most consistent associations were observed in Channels 23, 29, 35, 37, and 48. Some previous fNIRS studies reported that greater symptom severity was associated with reduced prefrontal activation. For example, Takizawa et al^[52]^. and Akiyama et al^[50]^. reported that reduced prefrontal and dorsolateral prefrontal activation during VFT was associated with greater depressive symptom severity. Importantly, our findings extend these studies by showing that dynamic hemodynamic features, particularly HbO Difference metrics, were stronger predictors of anxiety, depressive, and physiosomatic symptom severity than conventional activation measures such as β₁ or peak HbO.

Another novel finding is that PHQ-15 scores were significantly positively correlated with the connectivity strength between the right frontal pole (channel 36, MNI: 24, 72, 9) and the midline mPFC (channel 28, MNI: 1, 63, 28). The right frontal pole plays an important role in somatosensory integration, pain perception, and body representation^[65,66]^, whereas the midline mPFC is involved in interoceptive signal processing^[67]^. Therefore, increased connectivity in this pathway may reflect excessive attention to or misinterpretation of interoceptive signals in patients with AD and CO, manifesting as somatic symptoms. These findings support the concept physiosomatic symptoms, such as fatigue and pain, arise from the same disturbances in neurovascular coupling and cortical network organization that underlie anxiety and depression. This finding may provide a novel network-level perspective for understanding the neural mechanisms of somatization in ANSD.

## Limitations

Several limitations should be acknowledged. First, fNIRS measures cortical surface hemodynamics and cannot directly assess subcortical structures. Second, the cross-sectional design precludes causal inference; longitudinal studies are needed to firmly determine whether the identified markers reflect vulnerability, current state, or residual trait abnormalities. Third, although medication status was included in sensitivity analyses, treatment duration was not fully modeled. Fourth, the study was conducted at a single center in Chengdu, China and requires external validation in independent study cohorts. Fourth, although the VFT is a classic executive task, it captures limited emotion-specific processes. Future studies using emotional face or emotional Stroop paradigms may further reveal emotion-specific network alterations.

## Conclusions

This study demonstrates that ANSD are characterized by widespread abnormalities in task-evoked cortical hemodynamics, neurovascular coupling, functional connectivity, and network topology during verbal fluency processing. Collectively, the present findings are compatible with a state of neurovascular and microcirculatory dysfunction characterized by impaired cortical energetic efficiency and altered information processing across the anxiety spectrum. These findings may suggest peripheral vascular contributions to impaired neurovascular recruitment.

Our findings support a transdiagnostic model of ANSD, in which common neurovascular abnormalities cut across conventional diagnostic boundaries. Rather than being highly specific to GAD, AD, or CO, reduced prefrontal activation and increased hemodynamic instability are continuously associated with symptom severity across the entire spectrum, suggesting a shared underlying pathophysiological continuum. Consequently, the observed fNIRS abnormalities appear to reflect dimensional processes linked to overall illness severity, supporting a transdiagnostic neurovascular framework for ANSD psychopathology.

The observed combination of various hemodynamic aberrations in ANSD may reflect the impact of neuroimmune, metabolic, and oxidative stress (NIMETOX) pathways^[68]^ on the neurovascular unit. From a mechanistic perspective, endothelial dysfunction, autonomic dysregulation, oxidative stress, and inflammatory processes may impair nitric oxide-mediated vasodilation and microvascular reserve, thereby limiting cortical oxygen delivery under cognitive load. Immune-inflammatory activation and oxidative and nitrosative stress can impair endothelial function, astrocyte–vascular signaling, and nitric oxide-mediated vasodilation, thereby reducing task-evoked oxygen delivery and cerebral blood-volume recruitment^[15,68,69]^. In parallel, mitochondrial dysfunction may constrain neuronal energy production and neurovascular responsiveness, limiting cortical recruitment during cognitive demand^[70]^. Furthermore, autonomic dysregulation, a well-established feature of anxiety in affective disorders^[71]^, may increase short-term hemodynamic fluctuations and impair vascular adaptability, resulting in unstable neurovascular coupling.

## Supporting information

esf

## Data Availability

De-identified data and analysis code may be made available from the corresponding author (MM) upon reasonable request and subject to institutional approval.

## Abbreviations

AD: anxious depression
GAD: generalized anxiety disorder
CO: comorbid anxiety-depression
MDD: major depressive disorder
DLPFC: dorsolateral prefrontal cortex
FDR: false discovery rate
FEF: frontal eye fields
fNIRS: functional near-infrared spectroscopy
FPA: frontopolar area
GAD-7: Generalized Anxiety Disorder-7
GLM: general linear model
gPPI: generalized psychophysiological interaction
HC: healthy control
HbO: oxygenated hemoglobin
HbR: deoxygenated hemoglobin
HbT: total hemoglobin
OSOAS: Overall Severity of the Anxiety Spectrum
PHQ-9: Patient Health Questionnaire-9
PHQ-15: Patient Health Questionnaire-15
PLS-DA: partial least squares discriminant analysis
RF: random forest
ROI: region of interest
SMA: supplementary motor area
SVM: support vector machine
VFT: verbal fluency task.

## Declarations

### Ethics approval and consent to participate

The study protocol was approved by the Ethics Committee of Sichuan Provincial People’s Hospital [Approval No. 331 (2022)]. All participants or their legal representatives provided written informed consent before participation.

### Consent for publication

Not applicable.

### Authors’ contributions

Yiping Luo drafted the manuscript and contributed to interpretation of the findings. Hongzhou Wu performed the fNIRS analyses and prepared the figures. Xia Deng contributed to participant recruitment and clinical assessment. Luyao Wang and Yingqian Zhang and Andre F Carvalho contributed to data collection, data cleaning, and visualization. Xu Zhang served as co-corresponding author, provided clinical leadership, and supervised participant recruitment and clinical quality control and writing-review. Michael Maes served as corresponding author, supervised the study, contributed to conceptualization and interpretation, performed statistical analyses, and critically revised the manuscript. All authors read and approved of the final manuscript.

## Acknowledgements

None

