## Supplementary material for "Neurovascular instability, impaired cortical recruitment, and network dysconnectivity across the transdiagnostic anxiety spectrum: a functional multi-channel near-infrared spectroscopy study": esf

### Supplementary Materials

**Supplementary Table 1. Coverage of brain regions by 53 fNIRS channels.**

| Region of interest (ROI) | Channels |
|---|---|
| Left SMA | CH01 CH04 CH10 |
| Right SMA | CH40 CH47 CH52 |
| Left Broca's area | CH02 CH03 CH05 CH07 CH08 CH13 |
| Right Broca's area | CH44 CH46 CH49 CH50 CH51 CH53 |
| Left DLPFC | CH06 CH09 CH11 CH14 CH15 CH17 CH18 CH20 |
| Right DLPFC | CH31 CH32 CH34 CH39 CH42 CH43 CH45 CH48 |
| Left FEF | CH12 CH24 |
| Right FEF | CH26 CH38 |
| Left FPA | CH16 CH19 CH21 CH22 CH23 CH27 |
| Right FPA | CH30 CH33 CH35 CH36 CH37 CH41 |
| Midline | CH25 CH28 CH29 |

**Supplementary Table 2. FDR-significant differences in HbR and HbT hemodynamic features between the combined ANSD group and healthy controls.**

| Chromophore | Measurement | Region of interest (ROI) | Comparison | Adjusted mean diff | SE | 95% CI | p | FDR q |
|---|---|---|---|---|---|---|---|---|
| HbR | Difference | DLPFC_L | ANSD vs HC | 0.00092 | 0.00023 | 0.00047 to 0.00137 | <0.001 | <0.001 |
| | | DLPFC_R | ANSD vs HC | 0.00070 | 0.00018 | 0.00035 to 0.00105 | <0.001 | <0.001 |
| | | Broca_R | ANSD vs HC | 0.00051 | 0.00020 | 0.00011 to 0.00091 | .014 | .023 |
| | | SMA_L | ANSD vs HC | 0.00347 | 0.00067 | 0.00215 to 0.00480 | <0.001 | <0.001 |
| | | SMA_R | ANSD vs HC | 0.00124 | 0.00045 | 0.00035 to 0.00214 | .007 | .013 |
| | | FEF_R | ANSD vs HC | 0.00246 | 0.00066 | 0.00115 to 0.00376 | <0.001 | <0.001 |
| HbT | Difference | DLPFC_L | ANSD vs HC | 0.00080 | 0.00033 | 0.00015 to 0.00145 | .016 | .022 |
| | | DLPFC_R | ANSD vs HC | 0.00079 | 0.00025 | 0.00029 to 0.00129 | .002 | .008 |
| | | FPA_R | ANSD vs HC | -0.00046 | 0.00019 | -0.00083 to -0.00009 | .015 | .022 |
| | | Broca_R | ANSD vs HC | 0.00074 | 0.00026 | 0.00022 to 0.00125 | .005 | .013 |
| | | SMA_L | ANSD vs HC | 0.00209 | 0.00068 | 0.00075 to 0.00343 | .002 | .008 |
| | | SMA_R | ANSD vs HC | 0.00134 | 0.00053 | 0.00029 to 0.00240 | .013 | .022 |
| | | FEF_R | ANSD vs HC | 0.00443 | 0.00095 | 0.00256 to 0.00630 | <0.001 | <0.001 |
| | Peak | DLPFC_L | ANSD vs HC | -0.0901 | 0.0270 | -0.1435 to -0.0367 | .001 | .002 |
| | | DLPFC_R | ANSD vs HC | -0.0919 | 0.0276 | -0.1465 to -0.0374 | .001 | .002 |
| | | FPA_L | ANSD vs HC | -0.1256 | 0.0311 | -0.1869 to -0.0642 | <0.001 | <0.001 |
| | | FPA_R | ANSD vs HC | -0.1169 | 0.0342 | -0.1845 to -0.0494 | <0.001 | .002 |
| | | Broca_L | ANSD vs HC | -0.1120 | 0.0330 | -0.1772 to -0.0469 | <0.001 | .002 |
| | | Broca_R | ANSD vs HC | -0.1290 | 0.0357 | -0.1994 to -0.0585 | <0.001 | .002 |

Note: Results are covariate-adjusted estimated marginal mean comparisons from general linear models comparing the combined ANSD group (n = 96) with HC (n = 62), controlling for sex, age, and years of education. Mean difference was calculated as ANSD minus HC; positive values indicate higher values in ANSD, whereas negative values indicate higher values in HC. Pairwise comparisons used the least significant difference procedure and therefore yielded unadjusted p values. Benjamini-Hochberg FDR-adjusted q values were subsequently calculated across the 10 ROI-specific group comparisons separately within each chromophore-by-measurement family; only results with $q < 0.05$ are shown. Units: Difference, unitless; Peak and Mean, µmol/L; Integral, µmol·s/L. Abbreviations: ANSD, anxiety spectrum disorders; HC, healthy controls; HbR, deoxygenated hemoglobin; HbT, total hemoglobin; ROI, region of interest; SE, standard error; CI, confidence interval; FDR, false discovery rate; q, FDR-adjusted p value; DLPFC, dorsolateral prefrontal cortex; FPA, frontopolar area; SMA, supplementary motor area; FEF, frontal eye field.

**Supplementary Table 3. FDR-significant omnibus tests and post-hoc pairwise comparisons of peak and difference measures among the three ANSD groups.**

#### A. Covariate-adjusted omnibus tests surviving FDR correction

| Chromophore | Measurement | ROI | F | df1 | df2 | p | FDR q |
|---|---|---|---|---|---|---|---|
| HbO | Difference | DLPFC_L | 37.409 | 2 | 90 | 1.50E-12 | 3.00E-11 |
| HbO | Difference | DLPFC_R | 19.860 | 2 | 90 | 7.17E-08 | 7.17E-07 |
| HbO | Difference | Broca_R | 5.101 | 2 | 90 | .008 | .040 |
| HbO | Difference | SMA_R | 9.663 | 2 | 90 | 1.58E-04 | .001 |
| HbR | Difference | DLPFC_L | 65.880 | 2 | 90 | 2.38E-18 | 2.38E-17 |
| HbR | Difference | DLPFC_R | 22.309 | 2 | 90 | 1.35E-08 | 6.77E-08 |
| HbR | Difference | Broca_R | 6.533 | 2 | 90 | .002 | .006 |
| HbR | Difference | SMA_R | 13.144 | 2 | 90 | 9.82E-06 | 3.27E-05 |
| HbT | Difference | DLPFC_L | 31.669 | 2 | 90 | 3.86E-11 | 7.72E-10 |
| HbT | Difference | DLPFC_R | 15.515 | 2 | 90 | 1.62E-06 | 1.62E-05 |
| HbT | Difference | SMA_R | 6.360 | 2 | 90 | .003 | .017 |

B. Post-hoc pairwise comparisons surviving FDR correction

| Chromophore | Measurement | ROI | Comparison | Adjusted mean diff | SE | 95% CI | Post-hoc FDR q |
|---|---|---|---|---|---|---|---|
| HbO | Difference | DLPFC_L | CO vs AD | -0.00464 | 0.00059 | -0.00581 to -0.00347 | 8.12E-11 |
| HbO | Difference | DLPFC_L | AD vs GAD | 0.00440 | 0.00063 | 0.00316 to 0.00565 | 2.61E-09 |
| HbO | Difference | DLPFC_R | CO vs AD | -0.00290 | 0.00053 | -0.00395 to -0.00184 | 1.48E-06 |
| HbO | Difference | DLPFC_R | AD vs GAD | 0.00310 | 0.00057 | 0.00197 to 0.00422 | 1.48E-06 |
| HbO | Difference | Broca_R | CO vs AD | -0.00149 | 0.00062 | -0.00271 to -0.00026 | .027 |
| HbO | Difference | Broca_R | AD vs GAD | 0.00197 | 0.00065 | 0.00067 to 0.00327 | .005 |
| HbO | Difference | SMA_R | CO vs AD | -0.00423 | 0.00111 | -0.00644 to -0.00201 | 5.57E-04 |
| HbO | Difference | SMA_R | AD vs GAD | 0.00453 | 0.00119 | 0.00217 to 0.00688 | 5.46E-04 |
| HbR | Difference | DLPFC_L | CO vs AD | -0.00276 | 0.00027 | -0.00331 to -0.00222 | 6.43E-15 |
| HbR | Difference | DLPFC_L | AD vs GAD | 0.00285 | 0.00029 | 0.00227 to 0.00343 | 1.34E-14 |
| HbR | Difference | DLPFC_R | CO vs AD | -0.00163 | 0.00028 | -0.00218 to -0.00108 | 3.23E-07 |
| HbR | Difference | DLPFC_R | AD vs GAD | 0.00167 | 0.00029 | 0.00109 to 0.00225 | 6.89E-07 |
| HbR | Difference | Broca_R | CO vs AD | -0.00093 | 0.00036 | -0.00165 to -0.00021 | .019 |
| HbR | Difference | Broca_R | AD vs GAD | 0.00134 | 0.00039 | 0.00058 to 0.00211 | .001 |
| HbR | Difference | SMA_R | CO vs AD | -0.00324 | 0.00070 | -0.00463 to -0.00184 | 3.22E-05 |
| HbR | Difference | SMA_R | AD vs GAD | 0.00315 | 0.00075 | 0.00167 to 0.00463 | 1.34E-04 |
| HbT | Difference | DLPFC_L | CO vs AD | -0.00340 | 0.00047 | -0.00433 to -0.00247 | 1.15E-09 |
| HbT | Difference | DLPFC_L | AD vs GAD | 0.00322 | 0.00050 | 0.00223 to 0.00421 | 2.92E-08 |
| HbT | Difference | DLPFC_R | CO vs AD | -0.00195 | 0.00041 | -0.00277 to -0.00114 | 1.80E-05 |

| Chromophore | Measurement | ROI | Comparison | Adjusted mean diff | SE | 95% CI | Post-hoc FDR q |
|---|---|---|---|---|---|---|---|
| HbT | Difference | DLPFC_R | AD vs GAD | 0.00211 | 0.00043 | 0.00124 to 0.00297 | 1.54E-05 |
| HbT | Difference | SMA_R | CO vs AD | -0.00256 | 0.00084 | -0.00423 to -0.00089 | .005 |
| HbT | Difference | SMA_R | AD vs GAD | 0.00280 | 0.00089 | 0.00102 to 0.00458 | .004 |

Note: Omnibus tests were covariate-adjusted univariate general linear models comparing AD (n = 34), GAD (n = 28), and CO (n = 34), with HC excluded and sex, age, and years of education included as covariates. Benj amini-Hochberg FDR correction was applied separately across the 20 HbO tests, 10 HbR tests, and 20 HbT tests included in the Peak and Difference analysis; only omnibus effects with q < 0.05 are shown in panel A. Fo r outcomes with significant omnibus q values, panel B reports the prespecified post-hoc pairwise comparisons (GAD vs CO, CO vs AD, and AD vs GAD) that survived Benjamini-Hochberg FDR correction across all 33 post-hoc comparisons; non-significant post-hoc q values are not shown. Mean difference was calculated as the first group listed minus the second group; positive values indicate higher values in the first group. Unadjuste d post-hoc p values and omnibus q values are not reported in panel B; post-hoc q values are FDR-adjusted p values from the multiple pairwise comparisons. Units: Difference, unitless; Peak, µmol/L. Abbreviations: AD, anxious depression; GAD, generalized anxiety disorder; CO, comorbid anxiety and depression; HC, healthy controls; HbO, oxygenated hemoglobin; HbR, deoxygenated hemoglobin; HbT, total hemoglobin; ROI, region of interest; SE, standard error; CI, confidence interval; FDR, false discovery rate; q, FDR-adjusted p value; DLPFC, dorsolateral prefrontal cortex; FPA, frontopolar area; SMA, supplementary motor area; FEF, frontal ey e field.

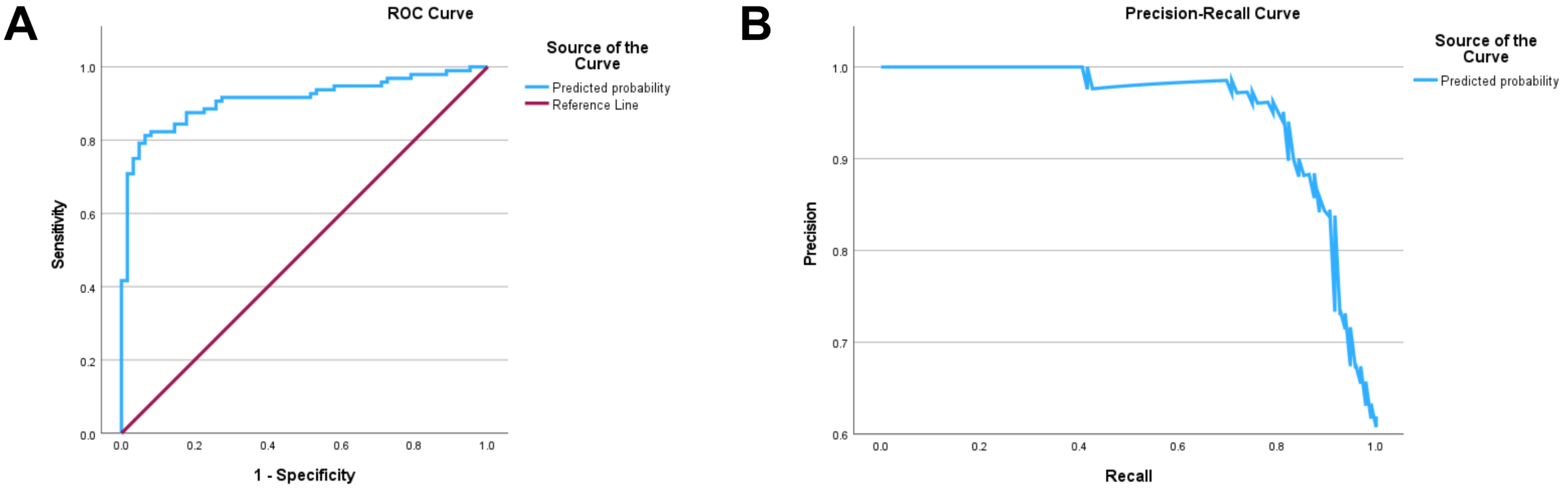


**Supplementary** Figure 1. Performance evaluation of the logistic regression model for predicting anxiety spectrum disorders status. (A) Receiver operating characteristic (ROC) curve, with an area under the curve (AUC) of 0.911 (95% CI: 0.865–0.958). The diagonal reference line represents random guessing (AUC = 0.5). (B) Precision-recall (PR) curve illustrating the trade-off between precision (positive predictive value) and recall (sensitivity) across different thresholds.

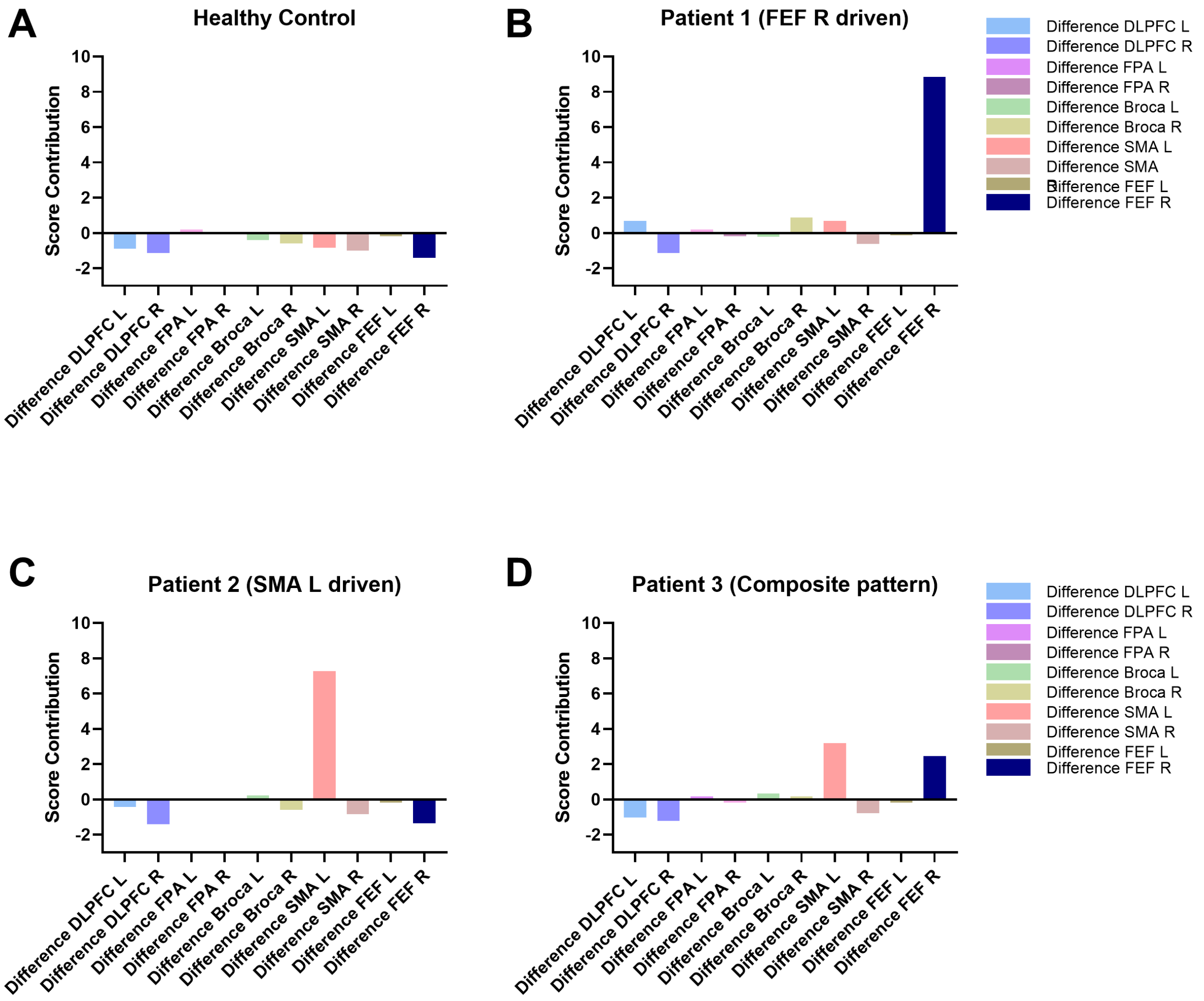


**Supplementary** Figure 2. Case-wise score contributions derived from the PLS-DA model for four representative participants. The charts illustrate the individualized contribution weights of the top 10 fNIRS features to the classification outcome. (A) A healthy control exhibiting generally negative or baseline contributions. (B-D) Three patients with anxiety spectrum disorders demonstrating distinct individual patterns: (B) predominantly driven by FEF_R, (C) driven by SMA_L, and (D) a combined pattern of FEF R and SMA L.

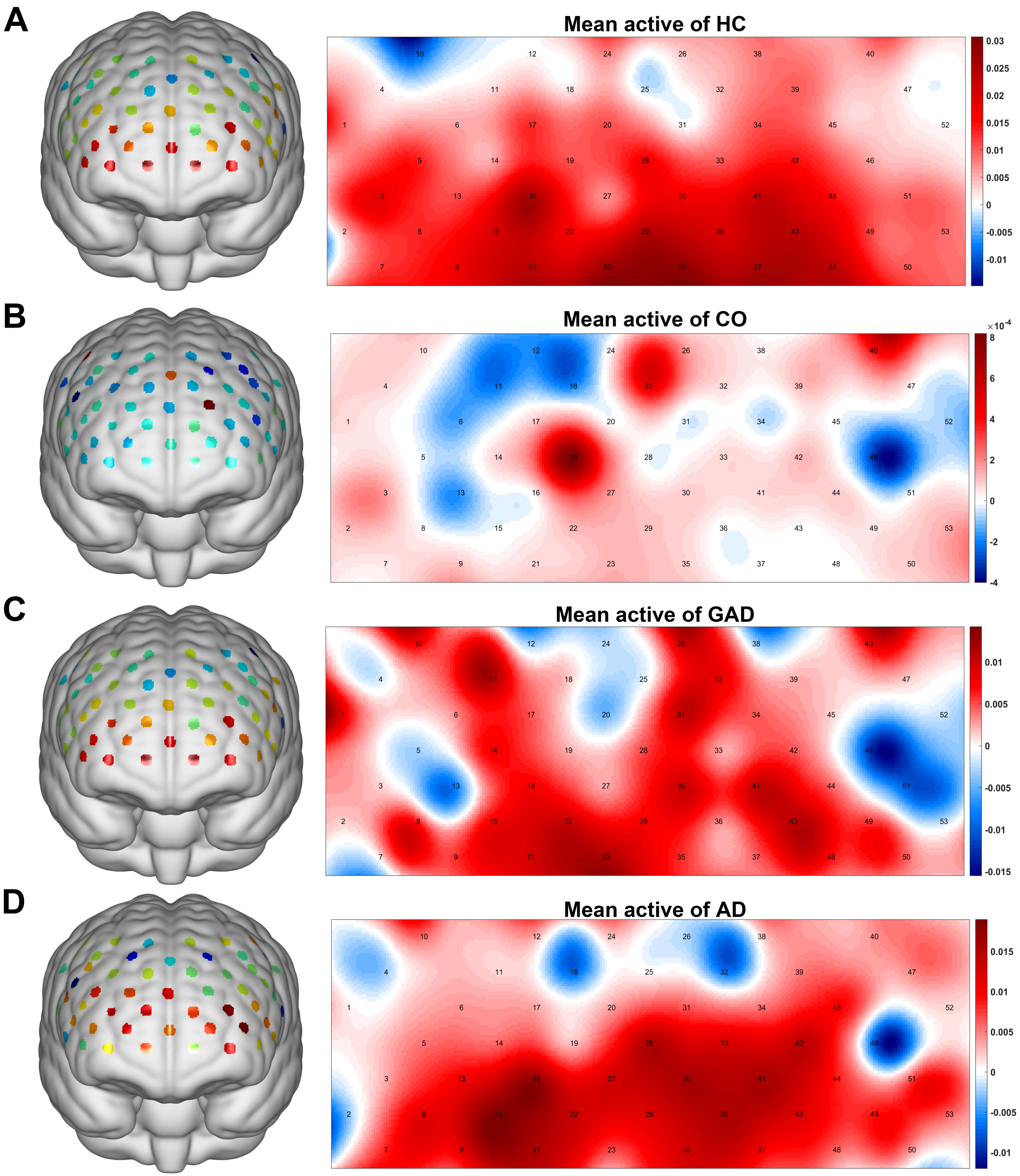
A
Mean active of HC
0.03
0.025
0.02
0.015
0.01
0.005
0
-0.005
-0.01
B
Mean active of CO
×10^-4
8
6
4
2
0
-2
-4
C
Mean active of GAD
0.01
0.005
0
-0.005
-0.01
-0.015
D
Mean active of AD
0.015
0.01
0.005
0
-0.005
-0.01

Supplementary Figure 3. Mean activation mapping of GLM analysis in the VFT task. (A)-(D) Mean activation mapping of the healthy control group, the comorbid anxiety-depression group, the generalized anxiety disorder, and the anxious depression group, respectively.
